# Integrated radiogenomics models predict response to neoadjuvant chemotherapy in high grade serous ovarian cancer

**DOI:** 10.1101/2021.07.22.21260982

**Authors:** Mireia Crispin-Ortuzar, Ramona Woitek, Elizabeth Moore, Marika Reinius, Lucian Beer, Vlad Bura, Leonardo Rundo, Cathal McCague, Stephan Ursprung, Lorena Escudero Sanchez, Paula Martin-Gonzalez, Florent Mouliere, Dineika Chandrananda, James Morris, Teodora Goranova, Anna M. Piskorz, Naveena Singh, Anju Sahdev, Roxana Pintican, Marta Zerunian, Helen Addley, Mercedes Jimenez-Linan, Florian Markowetz, Evis Sala, James D. Brenton

## Abstract

High grade serous ovarian cancer (HGSOC) is a highly heterogeneous disease that often presents at an advanced, metastatic state. The multi-scale complexity of HGSOC is a major obstacle to measuring response to neoadjuvant chemotherapy (NACT) and understanding its determinants. Here we propose a radiogenomic framework integrating clinical, radiomic, and blood-based biomarkers to measure and predict the response of HGSOC patients to NACT, showing how quantitative imaging data can serve as the backbone of multi-scale data integration. We developed and validated our approach in two independent highly-annotated multi-omic multi-lesion data sets. In a discovery cohort (n=72) we found that different tumour sites present distinct response patterns, and identified volumetric response assessment as a better predictor of overall survival (OS) than RECIST 1.1 status. We trained an ensemble machine learning approach to predict tumour volume response to NACT from data obtained prior to treatment, and validated the model in an internal hold-out cohort (n=20) and an independent external patient cohort (n=42). Benchmarking integrated models against models built on single data types highlighted the importance of comprehensive patient characterisation. Our study sets the foundation for developing new clinical trials of NACT in HGSOC.

## 1. Introduction

High grade serous ovarian cancer (HGSOC) remains a major therapeutic challenge and usually presents with advanced, multi-site metastatic disease. Neoadjuvant chemotherapy (NACT) followed by delayed primary surgery (DPS) is now the most frequent first line therapy for advanced HGSOC (1, 2). However, 39% of patients did not obtain any direct benefit from it in the large international ICON8 trial (3–5). Patient care would be substantially improved if these different sub-populations could be predicted before treatment started, for example by identifying likely non-responders who should receive immediate primary surgery. This variability in the response is driven by the complexity of HGSOC, which spans a large range of scales —from macroscopic tumour volumes observed on radiological imaging to microscopic immune microenvironments and sub-microscopic genomic diversity (6–8)—, but multiple-sampling strategies across different metastatic sites are not scalable.

Here we present an integrative radiogenomic framework to address the two fundamental challenges in the treatment of heterogeneous multi-site disease. The first challenge is accurately measuring response. The widely used RECIST 1.1 criteria are based on one-dimensional measurements performed on a small subset of lesions (9) and are thus insufficient for complex diseases such as HGSOC (10–12). This lack of accurate response measures prevents the optimisation of treatment strategies and the development of clinical trials for new combination therapies (13). The second major challenge is predicting response. Studies so far have focused on single data streams which can only offer a partial view of the disease, such as clinical features (14, 15), CA-125 (16–18), computed tomography (CT) imaging (19, 20), and circulating tumour DNA (ctDNA) (21). The superior predictive power of integrative models for complex endpoints is well documented in several cancer types (22–24), but training such models requires large, well-annotated, multi-omic datasets, which have not been available in HGSOC. In particular, radiological imaging is the only data source to capture the spatial heterogeneity of metastatic disease, but has so far been underused in HGSOC, where existing radiomics studies have focused only on correlations (19, 20), rather than combined predictive power, and none have considered NACT.

To overcome these challenges we used two independent highly annotated data sets to develop an integrated radiogenomic framework for measuring and predicting the response of HGSOC patients to NACT. Our framework showcases the power of quantitative imaging data to characterise metastatic disease and to serve as the backbone of multi-scale data integration.

## 2. Results

This study is based on two data sets, the first one (*n* = 92) from the NeOV trial and the second one (*n* = 42) from the Barts Health NHS Trust (Materials and Methods). Patients show large variability in treatment regimes and response patterns (Figure 1a). The NeOv dataset was randomly divided into a training set (*n* = 72) and a hold-out internal validation set (*n* = 20, Figure S1a). The training data set was used for the exploratory analyses and to train the machine learning models. The hold-out set was kept aside and only used for performance assessment after all predictive models had been fully trained. The Barts data were used as an independent, external validation data set.

**Fig. 1.**
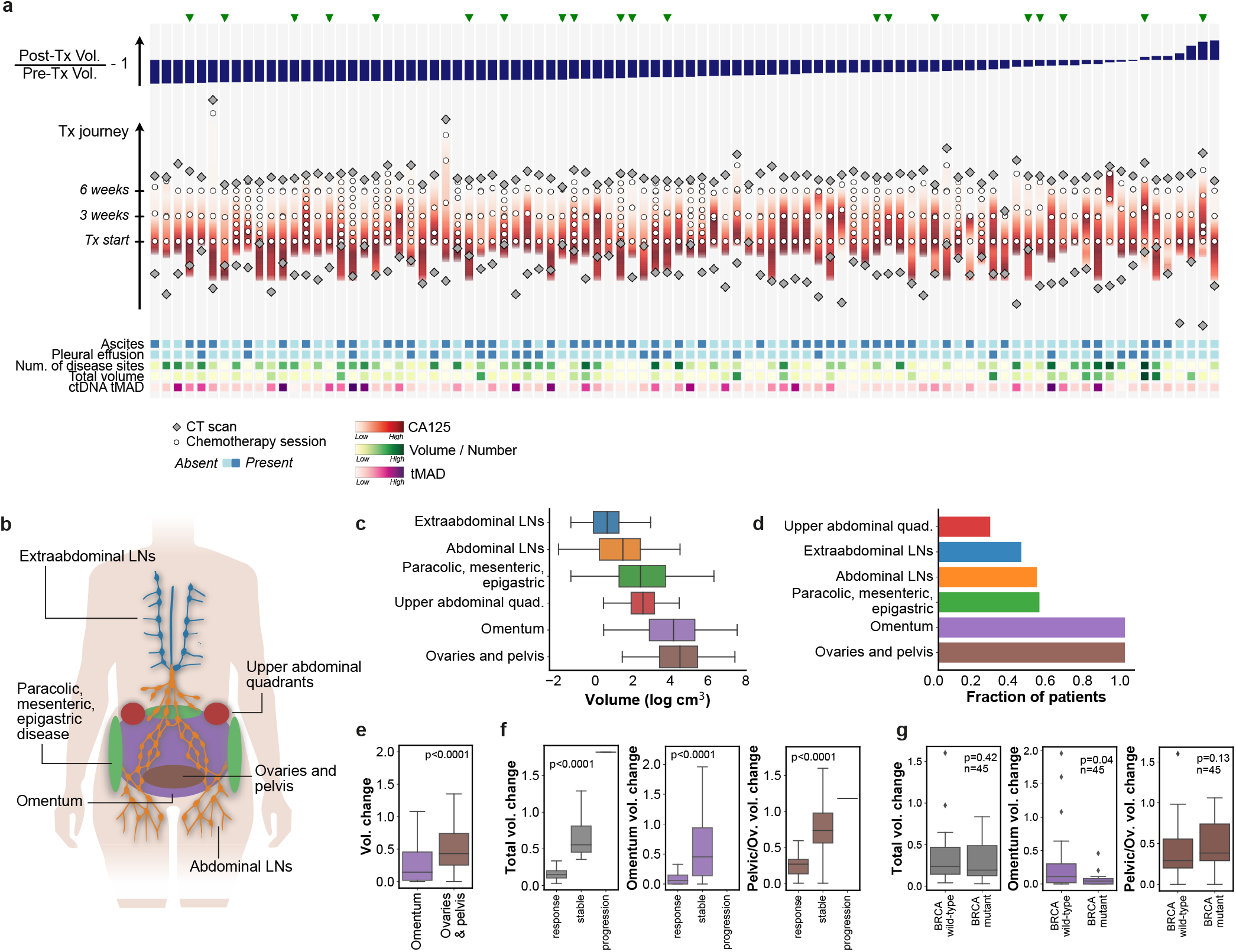
(a) Treatment courses of all 92 patients in the NeOV cohort, ordered by decreasing volumetric tumour response following NACT. Patients analysed in the hold-out validation set were randomly selected and are marked with a green triangle. Treatment journeys progress vertically (bottom to top) and are aligned at the time of the first chemotherapy session. Additional biomarkers obtained at baseline are depicted at the bottom as a heatmap. (b) Sites of primary and metastatic disease in HGSOC. (c) Distribution of the disease volumes found in the different sites for all the patients in the training cohort. (d) Fraction of patients in the training cohort with disease in the different sites. (e) Volume changes of the omental and pelvic/ovarian disease for all patients in the training cohort. (f) Total and site-specific volume change stratified by RECIST 1.1 response status. (g) Total and site-specific volume change stratified by *BRCA* mutation status. These figures are restricted to the n=45 patients in the training cohort for whom the *BRCA* mutation status was known. Boxes indicate the upper and lower quartiles, with a line at the median. Whiskers show the range of the data, and outliers are shown as circles and identified via the interquantile range rule.

### Response patterns to NACT are heterogeneous

All primary and metastatic lesions identified on pre- and post-NACT CT scans were segmented and labelled by a team of experienced radiologists (Figure 1b, see Materials and Methods). The omentum and the ovaries/pelvis were the two most frequent tumour locations (Figure 1d) and accounted for the majority of the disease burden at baseline (Figure 1c). The pattern of response differed between anatomical sites (Figure S2). Omental disease showed significantly better response than pelvic disease (Figure 1e). Most of that difference was explained by the particularly good omental response of patients with a *BRCA1/2* mutation (Figure 1g).

Despite the overall differences between sites, RECIST 1.1 status provided significant stratification for lesion-wise response (Figure 1f). In addition, the volume of omental disease at baseline was higher (*p* = 0.05) in responders as assessed by RECIST (complete or partial response; median = 85 cm^3^) compared to non-responders (stable disease or progression; median = 31 cm^3^).

While the number of disease locations at baseline was significantly correlated with response, disease volume at baseline (overall or in specific anatomic locations) was not (Table 1). This indicates that multivariable predictors are required to predict response to NACT rather than simple knowledge about disease burden and its anatomic distribution.

**Table 1.**
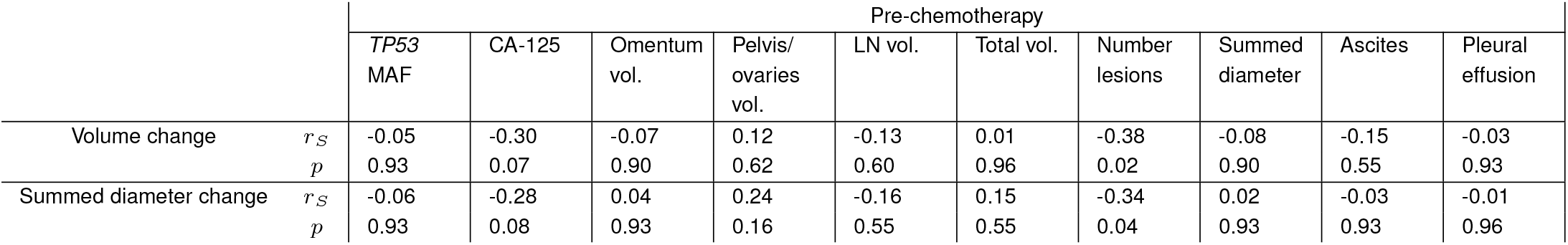
Spearman correlation coefficients between baseline measurements of tumour burden and different assessments of treatment response.

### ctDNA and CA-125 correlate with different types of disease burden

For all patients in the discovery cohort, ctDNA was assessed at baseline for the *TP53* mutant allele fraction (MAF), which has been proposed as a biomarker for HGSOC monitoring (25, 26). We also computed the trimmed median absolute deviation from copy number neutrality based on shallow whole genome sequencing (t-MAD), which has been shown to enhance tumour DNA detection (27). Owing to the strong correlation of t-MAD with *TP53* MAF (*p <* 0.0001), only *TP53* MAF was included in univariable analyses (Figure S1c).

Total disease burden at baseline (total volume, number of lesions, and summed RECIST 1.1 diameters) correlated significantly with both CA-125 and *TP53* MAF (Table 2). Neither baseline CA-125 nor *TP53* MAF correlated significantly with response (Table 1), but they did show a significant positive correlation with the summed RECIST 1.1 diameters post-chemotherapy.

**Table 2.** Spearman correlation coefficients between pre- and post-NACT measurements of tumour burden and blood biomarkers measured at baseline.

|  |  | Pre-chemotherapy |  |  |  |  |  |  |  |
| --- | --- | --- | --- | --- | --- | --- | --- | --- | --- |
|  |  | Omentum vol. | Pelvis/ ovaries vol. | LN vol. | Total vol. | Number lesions | Summed diameter | Ascites | Pleural effusion |
| Baseline *TP53* MAF | $r_S$ | 0.04 | 0.37 | 0.26 | 0.37 | 0.32 | 0.48 | 0.05 | 0.13 |
| | $p$ | 0.77 | 0.005 | 0.06 | 0.005 | 0.02 | 0.0003 | 0.77 | 0.36 |
| Baseline CA-125 | $r_S$ | 0.41 | 0.38 | 0.21 | 0.48 | 0.28 | 0.45 | 0.28 | 0.23 |
| | $p$ | 0.002 | 0.005 | 0.13 | 0.0003 | 0.04 | 0.0009 | 0.04 | 0.10 |

|  |  | Post-chemotherapy |  |  |  |  |  |  |  |
| --- | --- | --- | --- | --- | --- | --- | --- | --- | --- |
|  |  | Omentum vol. | Pelvis/ ovaries vol. | LN vol. | Total vol. | Number lesions | Summed diameter | Ascites | Pleural effusion |
| Baseline *TP53* MAF | $r_S$ | -0.04 | 0.30 | 0.13 | 0.29 | 0.19 | 0.41 | 0.03 | 0.13 |
| | $p$ | 0.79 | 0.03 | 0.36 | 0.03 | 0.17 | 0.003 | 0.79 | 0.36 |
| Baseline CA-125 | $r_S$ | 0.10 | 0.23 | 0.10 | 0.22 | 0.20 | 0.32 | -0.07 | -0.16 |
| | $p$ | 0.50 | 0.10 | 0.50 | 0.11 | 0.15 | 0.02 | 0.63 | 0.24 |

Baseline CA-125 correlated with both omental disease and pelvic and ovarian disease measured before chemotherapy. Similarly, baseline *TP53* MAF correlated significantly with pelvic and ovarian disease measured both before and after chemotherapy. However, it did not correlate with omental disease at either time point (Table 2). This suggests that high *TP53* MAF at baseline could be a specific indicator for high disease burden in the ovaries or pelvis, which tends to show poorer response (Figure 1).

### Some families of radiomics features correlate with clinical and biological characteristics

To capture the radiological complexity of the disease we defined several collections of radiomics features (see Table S9 for the full list). Volumes and number of lesions were calculated for the relevant anatomical sites. Shape features, first-order histogram statistics and texture features (‘intensity radiomics’) were calculated for each lesion and averaged over the whole disease. Intra-lesion heterogeneity was assessed by contracting the lesion contours and calculating the ratio of radiomics features before and after the contraction (‘rim radiomics’). Similarly, the external context of the lesions was assessed by calculating the ratio of radiomics features before and after dilating the contours (‘peripheric radiomics’). Finally, we also defined a series of binary variables to describe additional radiological findings (‘semantic features’): ascites and pleural effusion were assessed manually, and we used a previously developed automated tissue-specific sub-segmentation tool to identify hypodense (cystic/necrotic spaces) and hyperdense (calcifications) lesion parts within the manual segmentations (28).

We found that imaging features grouped into 6 distinct clusters (Figure 2). Cluster 1 was associated with baseline CA-125 levels (*r*_median_ = 0.34) and contained mostly lesion volume metrics. This is consistent with previous work suggesting that CA-125 correlates with lesion volume (29). Clusters associated with ctDNA features were generally dominated by features quantifying lesion heterogeneity and context. Cluster 5 was primarily associated with ctDNA *TP53* status (*r*_median_ = 0.25), and contained predominantly peripheric radiomics features, which quantify lesion context. Cluster 6 was associated with ctDNA *TP53* MAF and tMAD (*r*_median_ = 0.20 and 0.19 respectively), and contained predominantly rim ratio radiomics features, which provide information on intra-lesion heterogeneity.

**Fig. 2.**
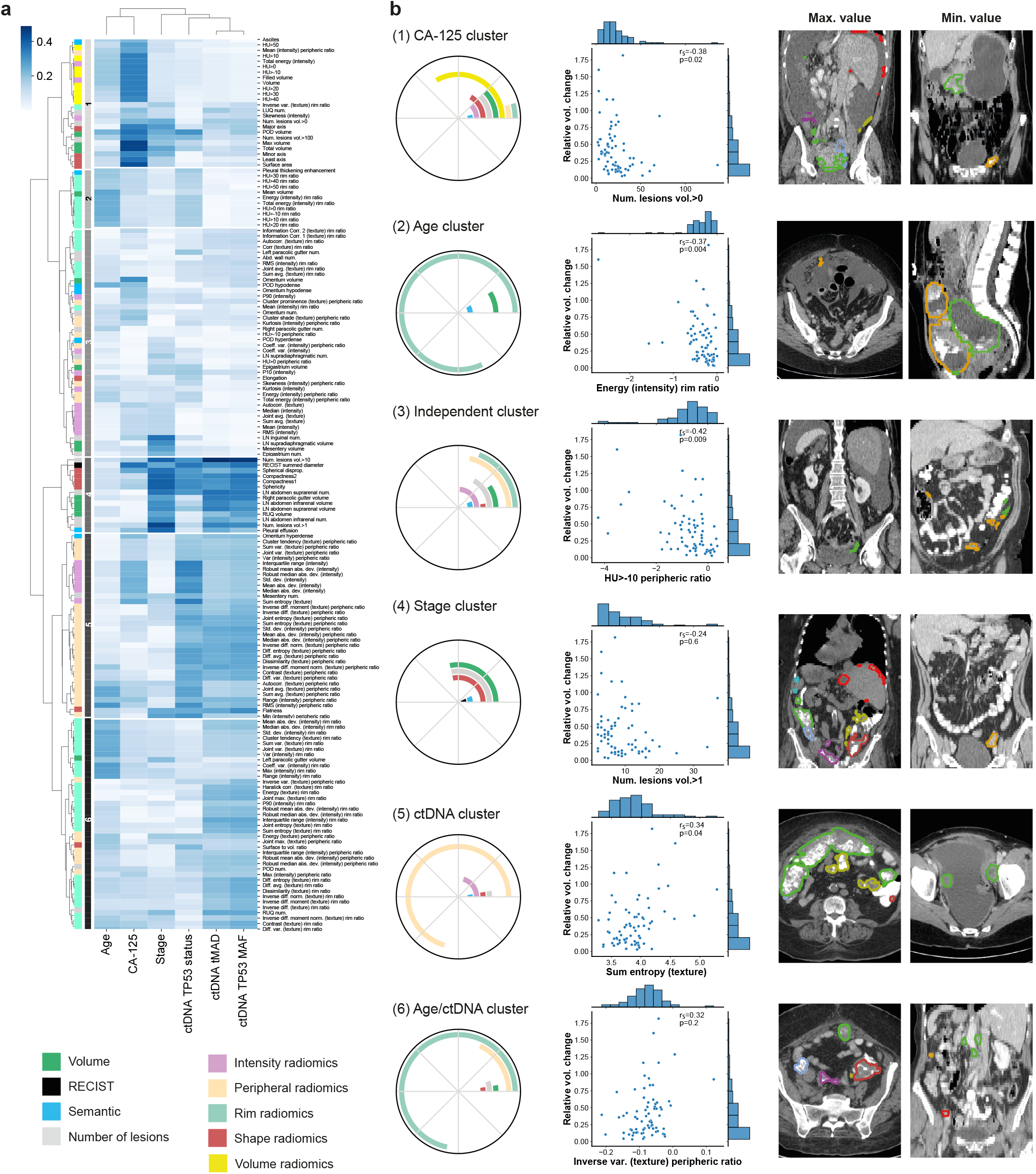
(a) Spearman correlation coefficients between imaging (rows) and clinical and biological features (columns), both clustered using a hierarchical approach. (b) Composition and characteristics of the six identified imaging feature clusters. Polar plots indicate the relative contribution of the different classes of imaging features. Scatter plots show the feature of each cluster with the highest Spearman correlation with volumetric treatment response. Each features is illustrated by displaying one slice from the patient with the maximum value (left), and one from the patient with the minimum value (right).

Cluster 4 was highly correlated to stage (*r*_median_ = 0.36), and was composed of a mixture of features related to shape, volume, and number of lesions, which quantify the disease burden. This is consistent with the definition of FIGO stage, which relies on the assessment of the extent and spread of the disease (30). Cluster 2 was associated mostly with age (*r*_median_ = 0.23) and contained almost exclusively rim ratio radiomics features, which provide information about intra-tumour heterogeneity. The remaining group (cluster 3) was formed by a heterogeneous mixture of features, and did not associate with any biological or clinical feature.

These results indicate that some of the information that global biomarkers such as stage, CA-125 or ctDNA provide can also be captured in multi-lesion radiomics features that quantify the extent, spread, heterogeneity and context of the disease. In addition, as shown in Figure 2, clusters 1, 2, 3, and 5 contain imaging features that are significantly correlated with volumetric response to treatment after multiple comparison correction. Crucially, the fact that cluster 3 has negligible biological or clinical associations suggests that there may be additional value in the integration of all sources of data to predict treatment response.

### Volumetric response is a better prognostic marker than RECIST 1.1 status

We considered three possible metrics for treatment response assessment: RECIST 1.1 response status, RECIST 1.1 summed diameter ratio, and total volume ratio. The three metrics have the advantage of being defined at the end of NACT, and can therefore be used as clinical trial endpoints for rapid response evaluation. The decision of which one to use for predictive modelling was based on the evaluation of their prognostic power. We built univariable Cox proportional hazards models for PFS and OS (Table S1). RECIST 1.1 status was not significant for either PFS or OS. PFS was significantly predicted by the summed diameter ratio (*p* = 0.014), with the total volume ratio (*p* = 0.077) showing a strong trend. OS was strongly predicted by the total volume ratio (*p* = 0.026) while the summed diameter ratio was borderline significant (*p* = 0.051).

We also studied the power to discriminate very good or very poor responders by stratifying Kaplan-Meier curves by the top and bottom 20th percentiles. We found that RECIST 1.1 status did not provide significant separation between the groups (Figure 3), for either PFS or OS. By contrast, top responders identified with the RECIST 1.1 summed diameter ratio (*p* = 0.03) and particularly the total volume ratio (*p* = 0.01) both had significantly longer PFS. Similarly, the worst responders identified according to the total volume ratio had significantly worse OS (*p* = 0.03), but no discrimination was provided by the ratio of summed diameters. We therefore decided to use the total volume ratio as the response metric on which to train our predictive modelling framework.

**Fig. 3.**
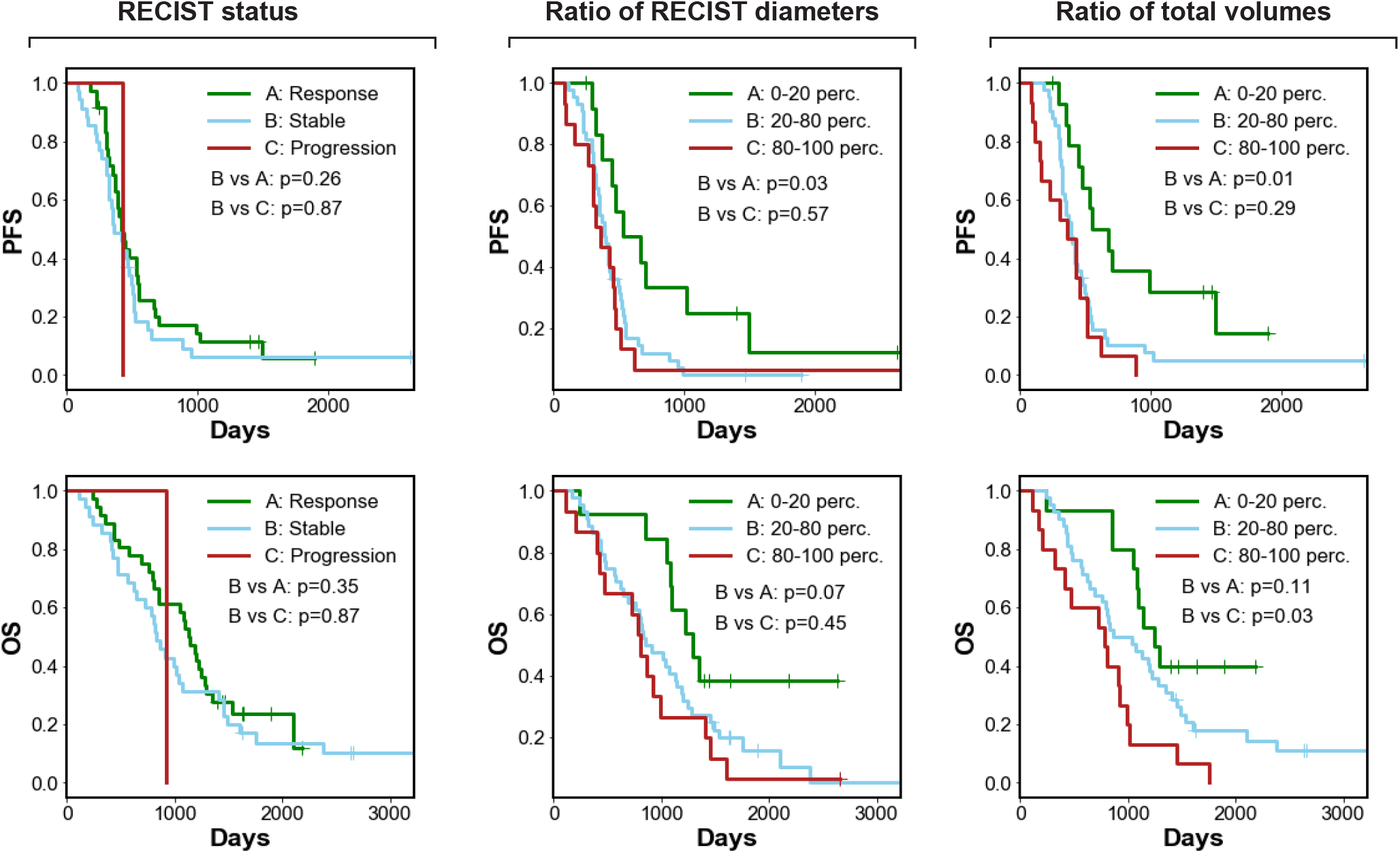
Kaplan-Meier curves for the three response metrics studied, from left to right: RECIST 1.1 status, RECIST 1.1 summed diameter ratio, and total volume ratio. The top row represents PFS, while the bottom row represents OS.

### Integrative models predict volume response to neoadjuvant chemotherapy

We built a machine learning framework to integrate the different streams of data into a set of predictive models of response to neoadjuvant chemotherapy. We used three distinct datasets in the process. The NeOv cohort (*n* = 92) was first randomly split into a training set (*n* = 72) and a hold-out validation set (*n* = 20). Model parameters were optimised and fixed using the training dataset in a cross-validation setup. The hold-out validation set was used as an independent test of model performance. A further external dataset, the Barts cohort, was used as an independent validation set (*n* = 42).

We trained a collection of models with an incremental, cumulative number of integrated features: age and FIGO stage, treatment characteristics, CA-125, radiomics features, and ctDNA, as depicted in Figure 4a. The full list of features is included in Tables S8 and S9.

**Fig. 4.**
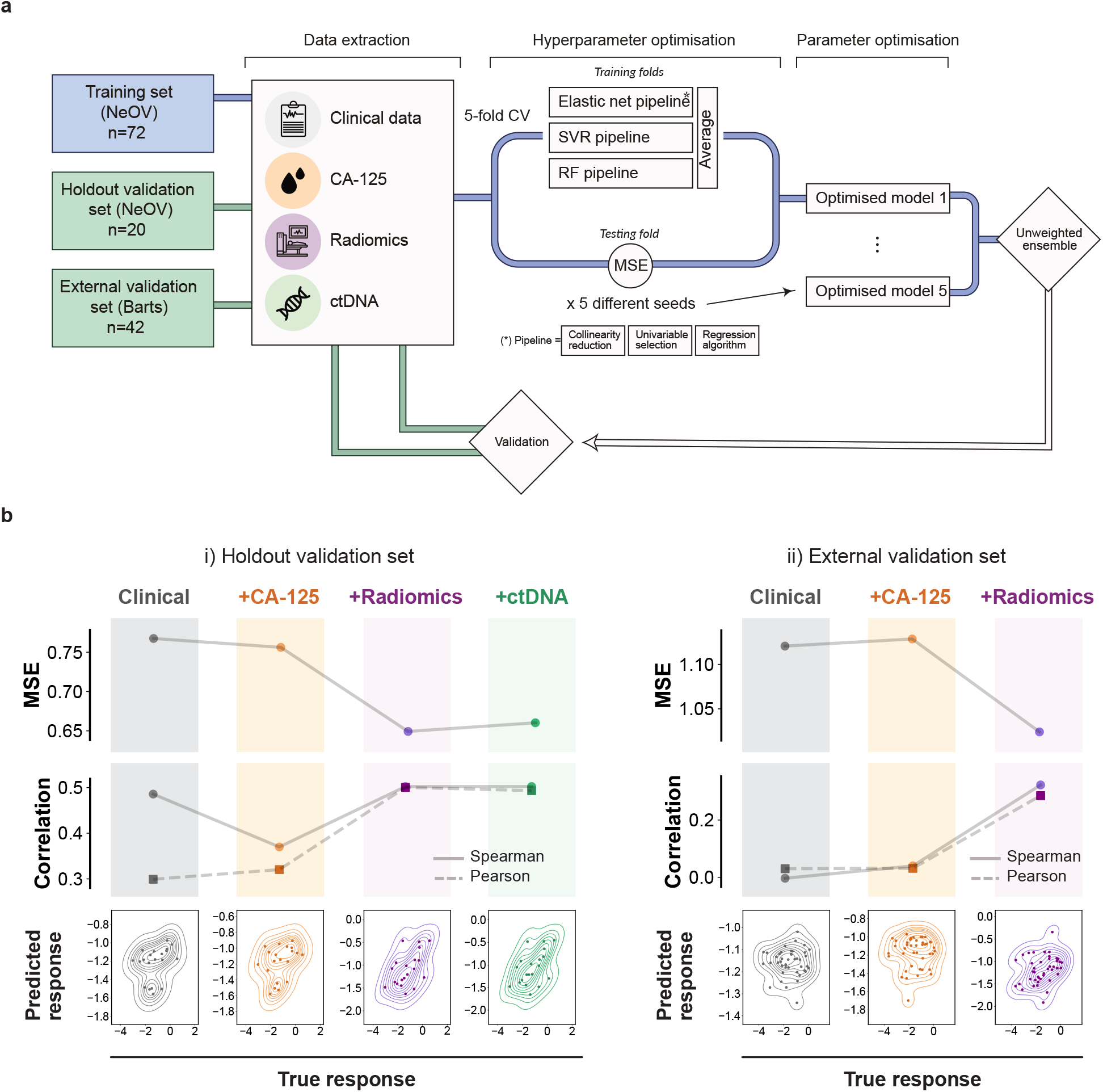
(a) Schematic of the machine learning framework for model training and validation. (b) Validation of the discriminative power of predictive models in the hold-out (left) and external validation cohorts (right), for models containing, from left to right, clinical, clinical+CA-125, clinical+CA-125+radiomics, and clinical+CA-125+radiomics+ctDNA features, respectively.

The response variable was defined as the logarithm of the post-chemotherapy total tumour volume divided by the pre-chemotherapy total tumour volume, as measured on the corresponding CT scans. This choice of response variable was motivated by the previous observation that volume shrinkage indicates longer OS and PFS (Figure 3), and is therefore prognostically important.

Model predictions were obtained by averaging the outputs of three machine learning pipelines based on elastic net, support vector regression, and random forest, respectively. The pipelines also included collinearity reduction and feature selection steps. We optimised model hyperparameters by minimizing the mean square error (MSE) in a 5-times 5-fold cross-validation scheme applied on the training cohort. During the training process, we found that the cross-validated MSE was reduced by 25% as a result of the successive integration of clinical, radiomic, and blood-based biomarkers. Adding ctDNA only resulted in a marginal improvement from 24% to 25% reduction in MSE (Table S7).

The final models were obtained by fixing the optimal hyperparameters and re-fitting to the entire training cohort. To assess the discriminative performance and calibration of the final models we applied them on the hold-out validation cohort. We observed a similar, gradual reduction in MSE, reaching a reduction of 15% for the integrated model without ctDNA, and 14% after integrating ctDNA (Figure 4 and Table S7). In addition, radiomics and ctDNA cumulative integration models produced response scores that were significantly correlated with the observed volume response (Spearman *r* = 0.5, *p* = 0.02 in both cases, Figure 4b and Table S7). Although the models were not trained to predict RECIST 1.1, we observed that the predicted scores were able to correctly rank the three RECIST 1.1 response groups in both the radiomics and ctDNA cumulative integration models (*p* = 0.02, Figure 5).

**Fig. 5.**
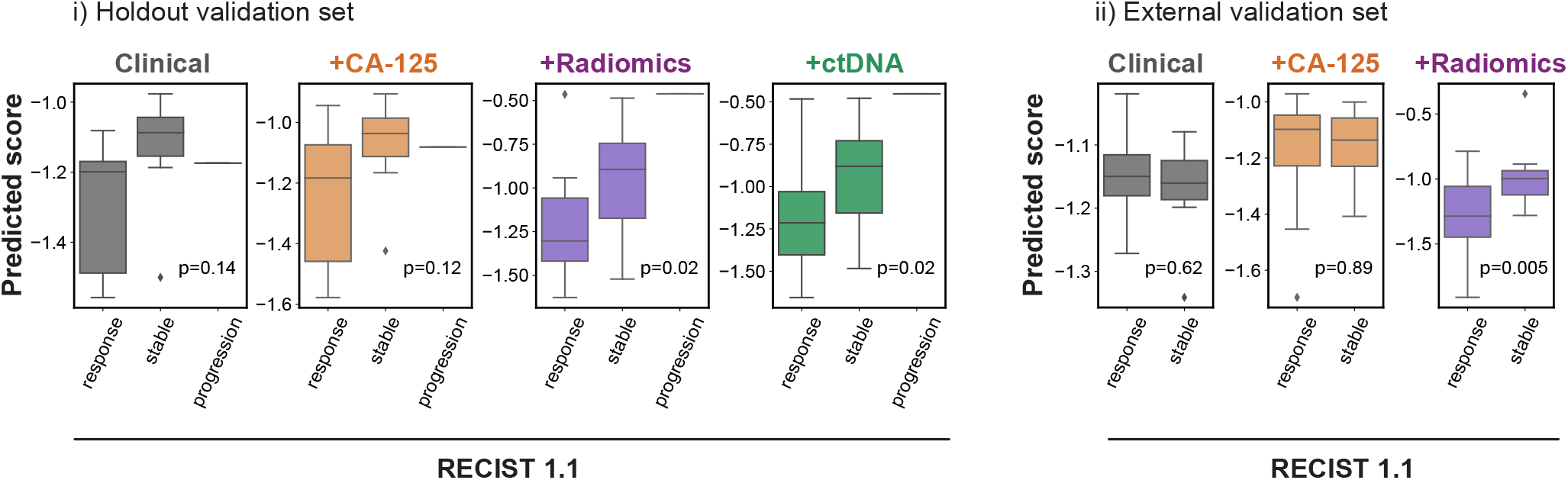
Validation of the ability of predictive models to describe RECIST 1.1 response in the hold-out (left) and external (right) validation cohorts, for models containing, respectively, clinical, clinical+CA-125, clinical+CA-125+radiomics, and clinical+CA-125+radiomics+ctDNA features. Boxes indicate the upper and lower quartiles, with a line at the median. Whiskers show the range of the data, and outliers are shown as circles and identified via the interquantile range rule.

We tested the generalizability of the models by applying them on an independent external cohort (Barts cohort, *n* = 42). As this cohort of patients did not have ctDNA data available, we were able to test the clinical, CA-125, and radiomics integration models only. We confirmed that integration was beneficial for discrimination, as only the radiomics integration model produced significant response scores (Spearman *r* = 0.32, *p* = 0.04, Figure 4b and Table S7), with a reduction in MSE of 8%. We also confirmed that the predicted scores were able to correctly rank the RECIST 1.1 response groups in the radiomics integration model (*p* = 0.005), but not the simpler ones (Figure 5).

We studied the relative contribution of the features used by the final models in two different ways. First, we examined which features passed the feature selection step in the three different component pipelines that integrate the machine learning models. Figure 6a shows the number of pipelines that selected a particular feature in each of the cumulative models, a metric that we call the ‘selection frequency’. We found that the treatment regimen (whether the patient received paclitaxel weekly, and whether the patient received carboplatin only) and the number of cycles of chemotherapy before the second scan were consistently selected across models. We also found that CA-125 was used in models that did not include radiomics, but was dropped from radiomics integration models. Semantic features, in particular pleural thickening and the presence of a hyperdense region in an omental lesion, were selected across all the relevant models. Mean volume, number of lesions, and volume and number of infrarenal lymph nodes were also selected. A small and consistent number of radiomics features were also selected, most of them belonging to the category of features defined to quantify lesion context.

**Fig. 6.**
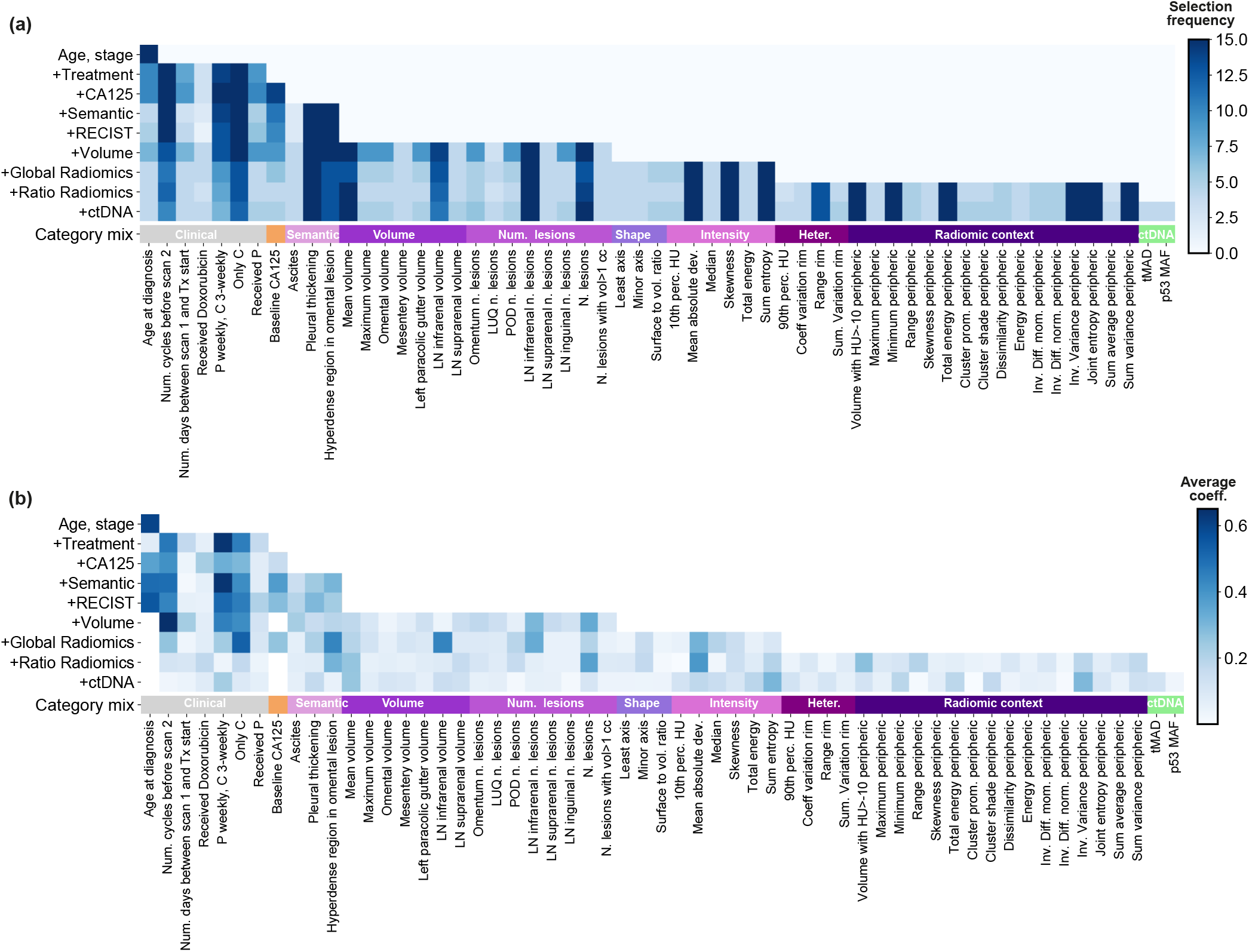
Importances of the features used by the predictive models. (a) Selection frequency. The heatmap shows the number of times that a given feature was selected in a model. The different rows correspond to different models with increasing, cumulative numbers of input features. As the optimisation is repeated 5 times, the range of the selection frequency is 0-15 (3 algorithms in the ensemble times 5 repetitions). (b) Averaged, normalised feature importances for the elastic net and random forest components of the models. Importances are defined from the feature coefficients for the elastic net regression, and from impurity-based Gini importances for random forest.

We also studied the feature importances in the elastic net and random forest component pipelines. To quantify the importances we used feature coefficients in the elastic net pipeline, and impurity-based Gini importances for the random forest pipeline. Figure 6b shows the averages of the normalised feature importances for those two component pipelines, for each of the 9 cumulative integration models. We found that most of the models integrating a large number of features tend to be more dense, with features sharing similar, lower importance levels. Features that tended to have larger importances were generally consistent with those that had the highest selection frequency, as can be observed by comparing the two panels in Figure 6.

## 3. Discussion

The clinical presentation of HGSOC is with complex, highly heterogeneous disease that is invariably metastatic throughout the abdomen. The accompanying genomic and cellular heterogeneity has impeded therapeutic progress and strongly suggests that understanding response to treatment must involve the integration of data from different sources and scales. To date, this has had not been systematically attempted. We have shown that integrated machine learning models based on baseline multi-scale data predict volumetric response to NACT (*p* = 0.04, external validation cohort). Our approach uses radiomics, the only data source capable of simultaneously measuring the entire metastatic disease, as the backbone of the prediction. Radiomics features had the strongest effects in the prediction models, which were validated in an independent cohort. We also found that volumetric response was a better predictor of OS (*p* = 0.03) than RECIST 1.1 status (*p* = 0.3) and revealed site-specific patterns of differential response and correlation with ctDNA detection.

The field of radiomics has grown exponentially in recent years, showing great promise across tumour sites and endpoints (31). At the same time, radiomics has been criticised for lack of robustness and reproducibility, as well as lack of biological interpretability (32, 33). Our study shows that both problems can be overcome by the right design choices. We made robustness a design priority for our predictive framework, which included strong feature selection, model ensembles on multiple levels, and repeated re-shufflings of the data to avoid biases. In addition, we trained the model on a dataset with heterogeneous imaging parameters (34); and we included new families of imaging features based on ratios between different volumes of interest, which were designed to partially cancel out such biases. We also curated an independent cohort from a different institution for validation, which we were able to perform successfully.

Our feature definitions were also designed to improve interpretability: the ratio features are not only robust to imaging parameters, but they also helped us to explore internal heterogeneity (rim features) and the external context of the lesions (peripheric features). Interestingly, we found that ratio features were the most numerous in the final models. This result is in line with previous work that found that qualitative radiological features describing the edges of peritoneal disease (nodular, diffuse, or mixed) were significantly associated with CLOVAR subtype (35) and BRCA mutation status (36). The concept of peritumoural radiomics has been explored before in breast, lung, and liver cancer (37–40), but to our knowledge our study is the first one to apply it to ovarian cancer. Our findings motivate the development of more detailed studies focusing on the boundary regions of ovarian cancer lesions – and show that data-driven radiomics analyses can support biological hypothesis generation.

Even if the hypothesis-motivated features had not played a significant role, the interpretability of the results was naturally supported by the integrative framework that the radiomics analysis was embedded in. For example, our study confirms previous observations that the presence of ctDNA is correlated with volume of disease at the start of treatment in HGSOC patients (26). The integration of clinical data into our models also yielded important insights, as features describing the type of NACT and its timing were consistently selected by our models. The ICON8 study (to which part of our discovery cohort was recruited) showed that PFS in patients with ovarian cancer undergoing NACT was unaffected by the administration regimen of paclitaxel (3-weekly as is standard or in a dose-dense weekly regimen) (41). However, the feature describing the mode of administration (weekly versus 3-weekly) was consistently selected by our prediction models. There are several possible explanations for this, including the limited size of our dataset, or a possible non-linear interplay with some of the other variables in the model. Alternatively, the administration mode could potentially be a factor that affects response to NACT but not survival. The median number of NACT cycles in our training cohort was lower (median of 3 cycles, with patients receiving weekly treatment having a median of 8 cycles) and may be more relevant for predicting response than survival. This is still clinically important, since disease extent after NACT affects resectability of the disease at DPS and therefore perioperative morbidity and operating time (42).

Addressing the challenges of reproducibility and interpretability is only the first step. Critically, previous studies have focused on a single ovarian lesion per patient as a proof of principle (19), without exploiting the full potential of imaging data to capture metastatic disease. Instead we enriched our data set by segmenting the full disease burden and including radiomics of all abdominopelvic tumour locations, reaching a median of 18 volumes of interest per scan in the training set. Our analysis showed that response to NACT in HGSOC varies spatially: lesions in the ovaries and/or pelvis had poorer response than those in the omentum. Ours is also the first study to integrate imaging features for all of the abdominopelvic disease with blood-based biomarkers, including ctDNA and CA-125. We showed that global tumour burden and volume of pelvic and ovarian tumors at baseline are significantly correlated with both CA-125 and ctDNA *TP53* MAF. However, our analysis of volumetric data also showed that the ctDNA signal was only due to the presence of disease in the ovaries and/or pelvis, suggesting that the simultaneous reading of ctDNA and CA-125 at baseline could play a role in helping determine disease spread and response in the diagnostic setting.

Some of these findings highlight the limitations of previous studies and ours. For example, even though we found that change in total volume may be a better prognostic indicator than RECIST-1.1, it is still treating the disease as a single entity, ignoring the subtle interplay between the pelvis/ovaries and the omentum. Similarly, the importance of some data streams is driven by the specific features that were included in the model. Indeed, given the correlation between ctDNA and total disease volume, and the higher dimensionality of the collections of clinical and radiomics features, it is not surprising that we did not find any additional value in adding ctDNA based biomarkers to our integrated models. However, our study was limited to ctDNA t-MAD, *TP53* MAF and *TP53* mutation status; a higher-dimensional panel of ctDNA sequencing data, applied on a larger dataset, and potentially measured longitudinally across different time points, may find that there are other complementarities between ctDNA and imaging data. Cell-of-origin analysis via mutation, methylation or fragmentomics to infer contribution from specific cell types to the circulating free DNA pool could help to predict specific volume reductions. Our dataset also lacked detailed quantification of *BRCA1/2* status and homologous recombination deficiency, which are likely to be important factors and should be incorporated into future models (43).

Another key limitation of the study is that manual segmentation for volumetry is still extremely time consuming. However, accurate and highly adaptable deep learning models for automatic segmentation are being rapidly developed across cancer types, facilitating the integration of volumetry into clinical workflows as well as large-scale radiomics computation (44).

In conclusion, our study is a proof-of-principle for the integration of multi-scale data to describe and predict the response of HGSOC patients to NACT. We demonstrate that the systematic multi-scale integration of standard-of-care biomarkers provides critical predictive power and important insights into the complex spatial configuration of the disease. After the necessary clinical validation in larger, prospective cohorts, a framework like the one we propose could have significant impact as a stratification tool in clinical and experimental settings –for example avoiding delays in surgery for patients who are unlikely to respond to chemotherapy–, and could bring forward a new generation of clinical trials for HGSOC, with rapid, effective endpoints that improve and expedite the discovery of new therapies.

## 4. Materials and Methods

### Patient cohorts

Two patient cohorts were used in this study. The main cohort (the ‘NeOV’ cohort) was randomly split into a training set and a hold-out set. The training set was used to train the machine learning models, and as a discovery dataset for univariable analyses. The hold-out set was set aside and used only to validate the model predictions. A second dataset (the ‘Barts’ cohort) was used for external validation.

For both data sets, patients had a confirmed histopathological diagnosis of HGSOC and were treated with neoadjuvant chemotherapy before delayed primary surgery. All patients within the main data set were treated at Cambridge University Hospitals NHS Foundation Trust between 2009 and 2020 and were recruited into a prospective clinical study approved by the local research ethics committee (REC reference numbers: 08/H0306/61). All patients within the Barts data set were treated at Barts Health NHS Trust between 2009 and 2018 and recruited into prospective clinical study approved by the local research ethics committee (IRAS reference numbers: 243824).

Written informed consent was obtained from all patients prior to any study related procedures. The study was performed in accordance with the principles of the 1964 Declaration of Helsinki and its later amendments or comparable ethical standards. Patients were identified and included based on the availability of at least two CT scans at baseline and prior to DPS. Additionally, for the main data set at least one baseline plasma sample for ctDNA assessments was required.

### Clinical data

Data regarding patient demographics, treatment, and disease were collected from the patient electronic medical records including notes from multidisciplinary team discussions (MDTs). PFS was defined as the time between histopathological diagnosis and first radiological evidence of progression or recurrence. Where progression date was unclear from radiology reports alone, (e.g. successive imaging studies with subtle/mixed changes), clinical interpretation of progression was incorporated in PFS date calling (e.g. documentation of breaking bad news to patients, treatment decisions for subsequent line therapy). OS was defined as the time from diagnosis to death. Stage was determined using the International Federation of Gynaecology and Obstetrics (FIGO) criteria for ovarian cancer (30).

### Management

The management of all patients in the study including indications for surgery were discussed and decided upon within MDTs as per the UK National Health Service (NHS) guidelines. Surgeries were performed through a midline laparotomy by a team specialised in surgical gynae-oncology aiming to achieve total macroscopic tumor clearance. Overall, *n* = 107 patients were treated with platinum-based chemotherapy in combination with paclitaxel, while 23 received carboplatin as a monotherapy. Patients were considered to have received weekly chemotherapy if the average time interval between doses ranged between 6 and 10 days, three-weekly if the average time interval was between 18 and 24 days, and irregular otherwise. Of those receiving combination therapy, 81 patients received carboplatin and paclitaxel three-weekly, 18 patients received carboplatin three-weekly and paclitaxel weekly, 2 patients received carboplatin and paclitaxel weekly, and the rest received the treatment at irregular intervals. In addition, 4 patients were treated with Doxorubicin. Table S2 shows the breakdown for the three datasets.

### BRCA status

Germline *BRCA1* and*BRCA2* mutational status was determined for 45 patients in the discovery cohort. The remaining cases reflect historical practice of not testing patients above 70.

### ctDNA

Blood samples were collected before initiation of treatment with chemotherapeutic agents. DNA was extracted from plasma (1.2-4ml) using QIAvac 24 Plus vacuum manifold and the QIAamp Circulating Nucleic Acid kit (Qiagen), or with QIAsymphony (Qiagen) as per manufacturer instructions. DNA quantification was performed using Qubit dsDNA broad-range or high-sensitivity assay kits and the Qubit Fluorometer (Thermo Fisher Scientific). Tagged-amplicon deep sequencing DNA libraries were prepared as described by Forshew et al. (45). Following purification with AMPure XP magnetic beads (Beckman Coulter Life Sciences), 10nM libraries were quantified using Agilent Bioanalyzer and Agilent DNA 1000 kit or Agilent TapeStation and ScreenTape D1000 (Agilent Technologies) according to manufacturer instructions, and pooled for sequencing on MiSeq, HiSeq 2500 or HiSeq 4000 (Illumina).

Shallow whole genome DNA libraries (10 million reads per sample) were prepared using the ThruPLEX DNA-Seq kit (Takara) and purified with AMPure XP magnetic beads (Beckman Coulter Life Sciences). 10nM libraries were quantified using Agilent D5000 ScreenTape System or Roche KAPA library quantification kits and pooled for sequencing on HiSeq 4000 (Illumina) in paired-end 150-base pair mode. On removing adapter sequences, shallow whole genome sequence reads were aligned to the 1000 Genomes Project version of the unmasked human reference genome GRCh37 using the BWA-MEM alignment software (46). Somatic copy number analysis was performed using CNAclinic (47) to generate trimmed Median Absolute Deviation from copy-number neutrality (t-MAD) scores as previously described (27).

Demultiplexed TAm-Seq reads were aligned to the GRCh37 reference genome by amplicon, and mutations called where non-reference alleles met probability criteria in both replicates, as previously described (45). Samples lacking mutation calls were manually curated using the Integrative Genomics Viewer (IGV)

### Imaging protocol

Clinically requested contrast-enhanced venous phase CT scans covering the abdomen and pelvis (with or without the chest depending on the clinical request and imaging findings) were either acquired at Cambridge University Hospitals NHS Foundation Trust (CUHNHSFT) or in other institutions across the UK and then imported into the picture archiving and communication system (PACS) at CUHNHSFT.

Therefore, different manufacturers and scanning protocols were used. Baseline scans were acquired between 0 and 14 weeks before initiation of neoadjuvant chemotherapy and post-treatment scans were acquired for response assessment after 1.6 to 5.8 months of treatment. All scans were initially identified on the local PACS and then fully anonymised for further study related processing.

## Data processing

### Image segmentation and labelling

On axial images reconstructed with a slice thickness of typically 5 mm (Table S3), and pixel spacings ranging between 0.053 and 0.095, using abdominal soft tissue window settings, all cancer lesions were segmented semi-automatically by a board certified radiologist with ten years of experience in clinical imaging, using Microsoft Radiomics (project InnerEye; Microsoft, Redmond, WA, USA). The volumes of interest (VOIs) were annotated for their anatomic location: omentum, right upper quadrant, left upper quadrant, epigastrium, mesentery, right paracolic gutter, left paracolic gutter, ovaries & pelvis, infrarenal abdominal lymph nodes, suprarenal abdominal lymph nodes, inguinal lymph nodes, supradiaphragmatic lymph nodes, other chest lymph nodes, parenchymal liver metastases, and lung metastases. Cystic and solid tumour parts were included in these segmentations. Automated sub-segmentation of hyperdense/calcified, hypodense/cystic or fatty and intermediately dense/solid tissue was performed for omental lesions and lesions of the ovaries and pelvis using a previously described and validated technique (28). Baseline and follow-up CT scans were evaluated according to RECIST 1.1 for response assessment (9). Pleural effusions and ascites were assessed semiquantitatively (0 = none, 1 = trace, 2 = less than 5 cm when measured perpendicularly to chest/abdominal wall, 3 = 5 cm or more when measured perpendicularly to chest/abdominal wall.

### Radiomics features

VOIs drawn manually were split into connected components using MATLAB’s *bwlabeln* function with a three-dimensional connectivity of 26, which assumes that voxels are connected if their faces, edges, or corners touch. Voxels with intensities below -100 HU were removed from the radiomics calculations. Radiomics features were extracted using the CERR Radiomics toolbox (48) (December 2018 version, GitHub hash: 5974376be7103d5c3831690c62aa721fc784d949), including shape, intensity-volume histogram, first-order, and Haralick texture features (see Table S9 for the full list). Intensity-volume histogram features, inspired by the Vx features commonly extrated in radiotherapy dose-volume histograms, corresponded to the volumes spanned by voxels above a certain intensity value (denoted ‘HU>x’ in Table S9). To calculate Haralick texture features for each lesion, co-occurrence matrices for 100 grey levels (up to a maximum of 1000 HU) were computed independently for each direction along 2D slices and averaged. To calculate the rim and peripheric radiomics features, for each VOI two copies were created by eroding and dilating the contours by 0.4 cm along the 2D slices. The value of the margin was chosen in order to capture slices of at least 1 cm diameter. Erosion and dilation were achieved by convolving the contour with a circular mask of the desired margin. Ratio features were computed by dividing the results obtained from the eroded and standard volumes (rim features), or the dilated and standard volumes (peripheric features). Shape features were not included in the ratios. Once standard and ratio features were calculated for each lesion, a single value was extracted for the whole patient by taking the unweighted mean of all lesions.

### Other imaging features

In contrast to the radiomics features, which we averaged across lesions, we did use the volume and the number of lesions in each of the anatomic locations as individual features. We also computed the mean, maximum, and total volume, as well as the number of lesions with volume bigger than 0, 1, 10, and 100 cm3. In addition, we defined four binary features that indicated whether or not there were hypodense or hyperdense regions in either omental or pelvic/ovarian lesions. Ascites and pleural effusion were used as defined by the radiologist, as explained above (section 4, Radiological image analysis).

### Clinical features

Chemotherapy regimens were extracted from the clinical records. We recorded whether the patient had received any Carboplatin, Paclitaxel or Doxorubicin in three binary variables. Mean periods were calculated by averaging the time intervals between sessions. We defined weekly regimen as having a mean period of 6 to 10 days, both included; and three-weekly as having a mean frequency of 18 to 24 days, both included. Typical combinations included weekly Paclitaxel and three-weekly Carboplatin; both weekly; and both three-weekly (Figure 1). These combinations were therefore encoded in binary variables. We also recorded whether patients had received Carboplatin only in a binary variable. FIGO stage was encoded by assigning ordinal numbers to in order of aggressiveness, from 1 for stage 1A to 10 for stage 4B. The exact mapping is listed in Table S4. CA-125 values were also extracted from the clinical records, with the measurement closest to the beginning of treatment being used for analysis. The full list of clinical and blood biomarker features can be found in Table S8.

## Statistical analysis

### Tumour burden correlations

Correlations were calculated using Spearman correlation coefficient and *p* values were corrected using the Benjamini-Hochberg procedure as implemented in the scikit-learn Python package (49). The total tumour burden affecting lymph nodes was calculating by combining the lesions found in infrarenal, suprarenal, inguinal, supradiaphragmatic, and chest lymph nodes. The difference between the relative volume change in the omentum and in the ovaries/pelvis was quantified using the Mann-Whitney U test. The difference in relative volume change for patients with different BRCA status was also quantified using the Mann-Whitney U test.

### Imaging clusters

To identify clinically or biologically meaningful clusters of radiomics features we clustered their Spearman correlation coefficients used a hierarchical clustering approach. The optimal number of imaging clusters was obtained by maximising their correlation with any of the clinical and biological features. To do this, we calculated the maximum Spearman correlation coefficient between each cluster and any of the biological/clinical features, and averaged the result across all clusters (Figure S3a). The metric reached a plateau at 6 clusters, which was therefore chosen as the optimal number (Figure S3b). The most predictive feature in each cluster (Figure 2) was chosen in terms of its Spearman correlation with volumetric treatment response. *p* values were corrected using the Benjamini-Hochberg procedure.

### Survival modelling

Cox Proportional Hazards modelling was performed with the Lifelines Python library (50). The baseline hazard was estimated non-parametrically using Breslow’s method. Parameter estimates, hazard ratios and 95% confidence intervals are listed in Table S1. Kaplan-Meier curves were also computed using the Lifelines library. 95% confidence intervals were computed using Greenwood’s Exponential formula. *p* values were computed using the log-rank test.

## Machine learning models

### Training

We created a machine learning framework to predict response to chemotherapy, evaluated on the basis of relative total volume change. We used the NeOv training set to train and optimise the models. Once trained and frozen, we evaluated the models in the internal hold-out set and in the external validation set. We used an increasing number of features, in order of general availability. We started with clinical features (age, stage, and treatment); then added baseline CA-125; then imaging features; and finally ctDNA. For each combination we retrained the framework and derived a new model.

The predictions were based on an unweighted ensemble regressor (51). The ensemble included three different machine learning algorithms: an elastic net, a support vector regressor with a radial basis function (RBF) kernel, and a random forest, all of them coded in Python using the scikit-learn package (49). Each algorithm was embedded in a scikit-learn pipeline with three pre-processing steps, namely collinearity reduction, z-score standardisation, and univariable feature selection. Collinearity reduction removed all features with a mutual Pearson correlation above 0.95, retaining only the one with the highest correlation with the response variable. The feature selection step removed all features that were not ranked within the top *k* according to their F-value. The scores produced by the three pipelines were averaged to form the prediction.

We used a 5-fold cross validation setup to optimise model hyperparameters in the training set, covering the hyperparameter ranges shown in Table S6. The optimisation was based on a randomised search in the hyperparameter space to optimise mean square error (MSE). Once the optimal hyperparameters were found, we determined model parameters by re-fitting the model to the entire training set. To increase model robustness, we repeated this process five times with five different cross-validation seeds. The five resulting optimal models were combined to form the final ensemble, in which the prediction is simply the average of the five predicted scores. In this regard, our ensemble setup has two different tiers: the randomisation tier (5 seeds), and the algorithmic tier (3 regressors acting in parallel for each seed).

### Validation

We validated the models on the hold-out internal validation set (*n* = 20) and the external validation set (*n* = 42). To quantify the calibration of the models, we computed the MSE. To quantify the discriminative power of the models, we computed the Spearman correlation coefficient and *p* value between the predicted and observed response scores. Finally, we evaluated whether the predicted scores were also able to rank patients into the different RECIST 1.1 categories using the *p* value associated with the point biserial correlation coefficient.

### Feature importance

We evaluated feature importances in two different steps. First we computed the frequency with which features were selected after the collinearity reduction and univariable selection steps. We repeated the process for each of the three algorithms and each of the five cross-validation seeds, which means that features could be selected between 0 and a maximum of 15 times, as seen in Figure 6. The table on Figure 6 displays only features that were chosen at least 3 out of 5 times in each cross-validation loops, for robustness. Second, we computed the importance of each individual feature within the regression algorithm. This was only possible for the elastic net, where we used the feature’s coefficients, and the random forests, where we used impurity-based feature importances. The results were averaged across the five seeds and the two algorithms, with the table on Figure 6 displaying only features that were chosen at least 3 out of 5 times in the cross-validation loops, as before.

### Data and code availability

The code and data can be found at https://github.com/micrisor/OvarianIntegration.

## Data Availability

The code and data can be found at https://github.com/micrisor/OvarianIntegration

## Acknowledgements

This work was partially supported by The Mark Foundation for Cancer Research and Cancer Research UK Cambridge Centre [C9685/A25177], the Wellcome Trust Innovator Award [RG98755] and the CRUK National Cancer Imaging Translational Accelerator (NCITA) [C42780/A27066]. Additional support was also provided by the National Institute of Health Research (NIHR) Cambridge Biomedical Research Centre (BRC-1215-20014). The views expressed are those of the authors and not necessarily those of the NHS, the NIHR or the Department of Health and Social Care. Microsoft Radiomics was provided to the Addenbrooke’s Hospital (Cambridge University Hospitals NHS Foundation Trust, Cambridge, UK) by the Microsoft InnerEye project.

## A. Supplementary figures

**Supplementary Fig. 1.**
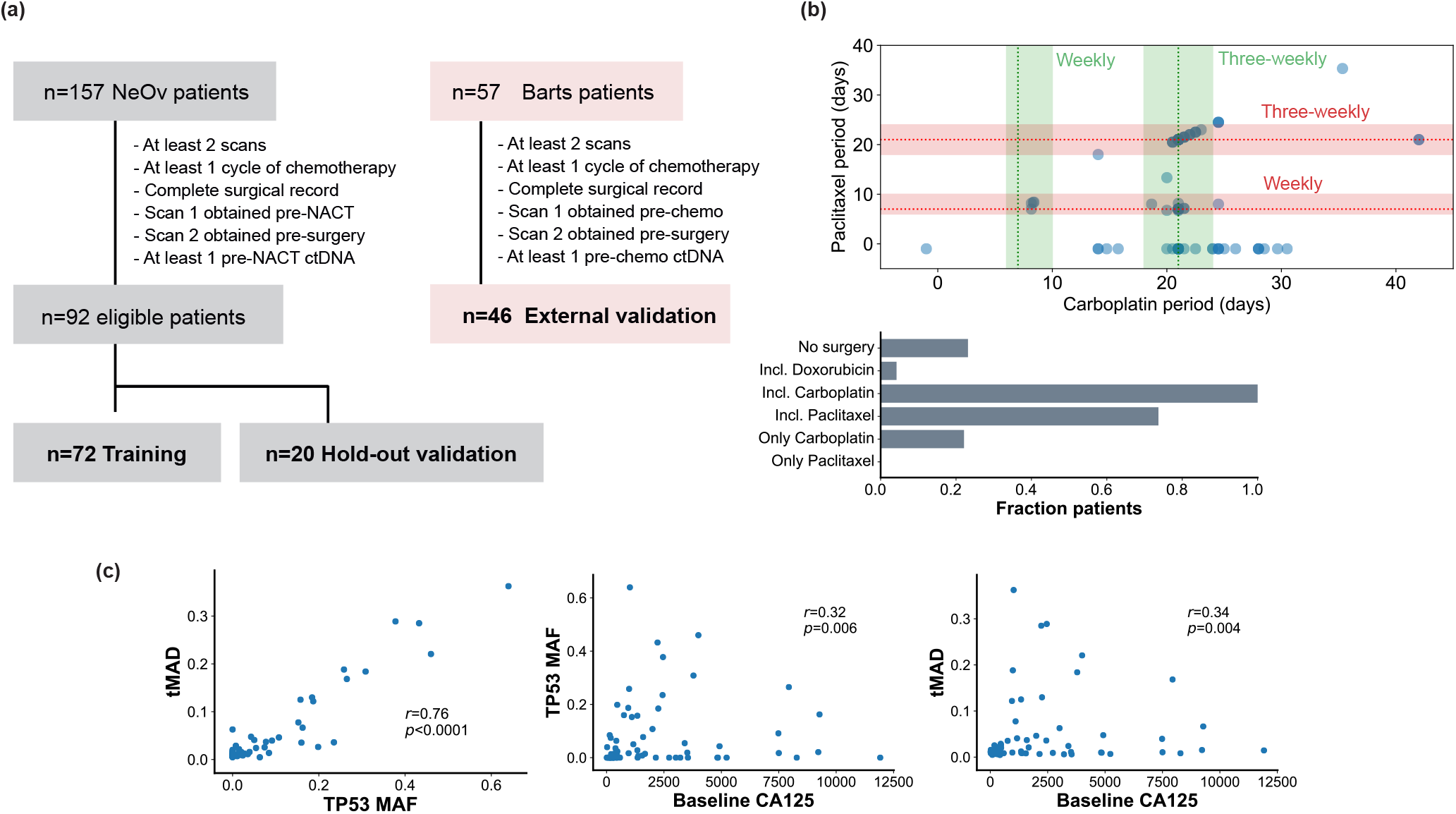
(a) Flowchart of the datasets used in the analysis. (b) Paclitaxel and carboplatin administration frequencies for patients in the NeOv cohort. Bands indicate the range of patients who are considered to have received either weekly, three-weekly, or other treatment frequency. (c) Scatter plots illustrating the correlations between the blood-based biomarkers.

**Supplementary Fig. 2.**
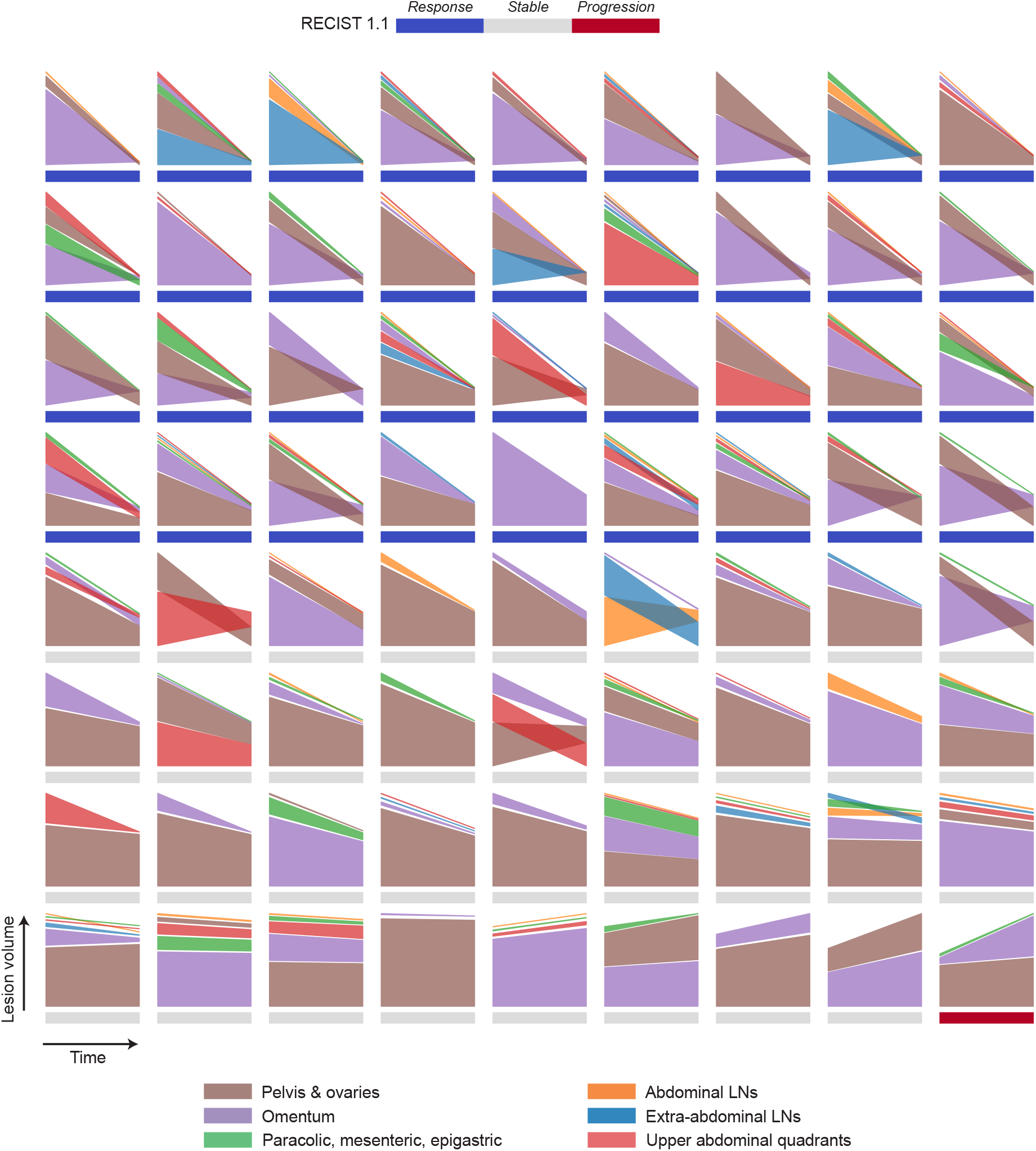
Site-specific change in volume for each of the patients in the training dataset. Patients are ordered as a function of the total change in volume.

**Supplementary Fig. 3.**
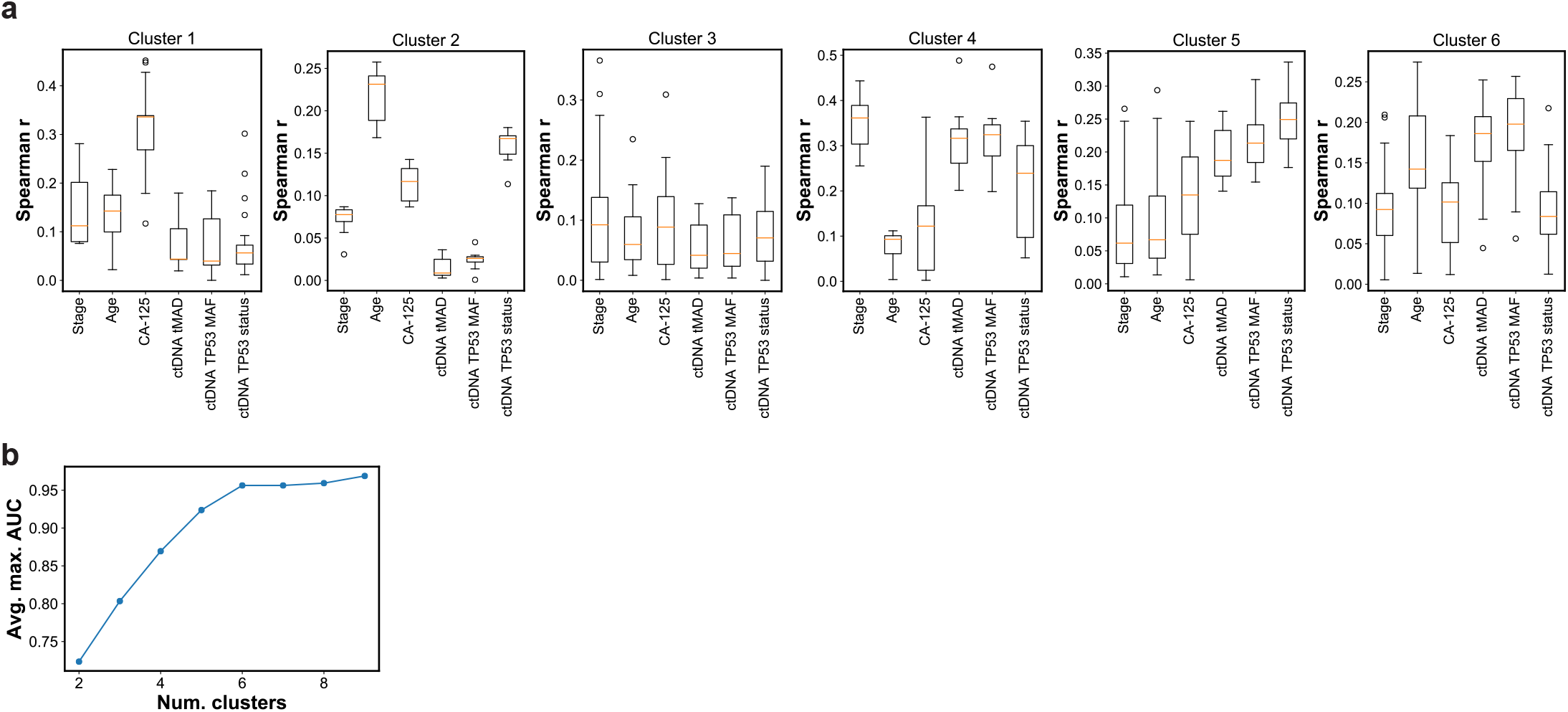
(a) Correlation between each of the imaging clusters and clinical features and blood biomarkers. Boxes indicate the upper and lower quartiles, with a line at the median. Whiskers show the range of the data, and outliers are shown as circles and identified via the interquantile range rule. (b) Optimisation of the number of imaging clusters.

## B. Supplementary tables

**Table 1.** Cox proportional hazards model results for overall survival (OS) and progression-free survival (PFS).

| Reference | Outcome | Parameter estimate | Standard error | HR | 95% CI | z-score | p-value |
| --- | --- | --- | --- | --- | --- | --- | --- |
| RECIST 1.1 | PFS | 0.2681 | 0.2347 | 1.3075 | -0.1920–0.7282 | 1.1420 | 0.2534 |
| RECIST 1.1 | OS | 0.2548 | 0.2474 | 1.2901 | -0.2301–0.7396 | 1.0299 | 0.3030 |
| Summed diameter change | PFS | 1.3414 | 0.5459 | 3.8244 | 0.2715–2.4113 | 2.4574 | 0.0140 |
| Summed diameter change | OS | 1.1173 | 0.5734 | 3.0567 | -0.0065–2.2411 | 1.9486 | 0.0513 |
| Volume change | PFS | 0.5540 | 0.3134 | 1.7402 | -0.0603–1.1684 | 1.7676 | 0.0771 |
| Volume change | OS | 0.7372 | 0.3308 | 2.0901 | 0.0889–1.3855 | 2.2287 | 0.0258 |

**Table 2.**
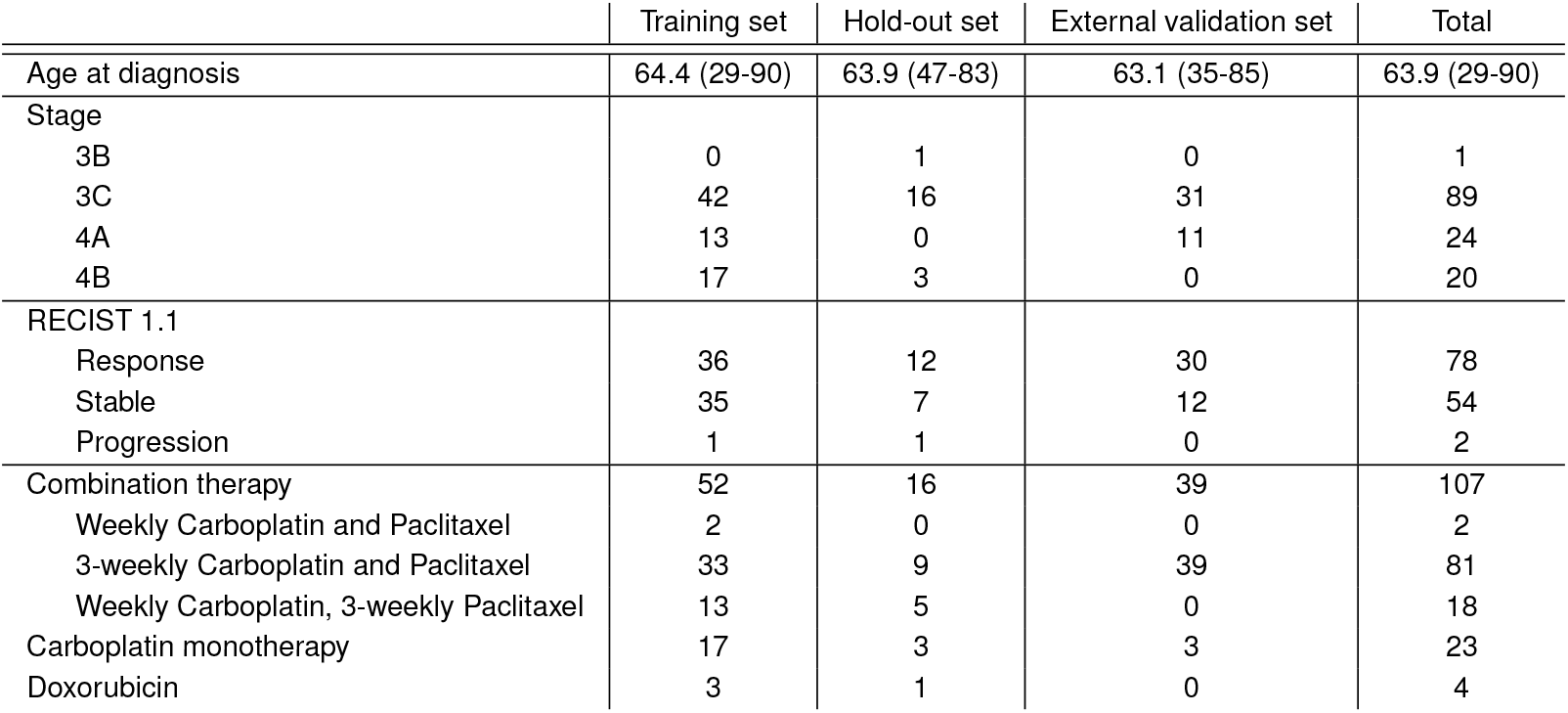
Breakdown of the age, stage, response status, and treatment regimens received by patients in the different cohorts.

|  | Training set | Hold-out set | External validation set | Total |
| --- | --- | --- | --- | --- |
| Age at diagnosis | 64.4 (29-90) | 63.9 (47-83) | 63.1 (35-85) | 63.9 (29-90) |
| Stage |  |  |  |  |
| 3B | 0 | 1 | 0 | 1 |
| 3C | 42 | 16 | 31 | 89 |
| 4A | 13 | 0 | 11 | 24 |
| 4B | 17 | 3 | 0 | 20 |
| RECIST 1.1 |  |  |  |  |
| Response | 36 | 12 | 30 | 78 |
| Stable | 35 | 7 | 12 | 54 |
| Progression | 1 | 1 | 0 | 2 |
| Combination therapy | 52 | 16 | 39 | 107 |
| Weekly Carboplatin and Paclitaxel | 2 | 0 | 0 | 2 |
| 3-weekly Carboplatin and Paclitaxel | 33 | 9 | 39 | 81 |
| Weekly Carboplatin, 3-weekly Paclitaxel | 13 | 5 | 0 | 18 |
| Carboplatin monotherapy | 17 | 3 | 3 | 23 |
| Doxorubicin | 3 | 1 | 0 | 4 |

**Table 3.** Slice thickness of the CT scans in the training and validation datasets.

| Dataset | Slice spacing [cm] (number of scans) |
| --- | --- |
| Training set | 0.069 (1), 0.15 (1), 0.25 (1), 0.3 (2), 0.375 (19), 0.5 (48) |
| Hold-out validation set | 0.3 (1), 0.375 (4), 0.5 (15) |
| External validation set | 0.069 (1), 0.2 (12), 0.41 (2), 0.5 (31) |

**Table 4.** Mapping between FIGO stages and ordinal numbers used for predictive modelling.

| FIGO stage | Ordinal |
| --- | --- |
| 1A | 1 |
| 1B | 2 |
| 1C | 3 |
| 2A | 4 |
| 2B | 5 |
| 3A | 6 |
| 3B | 7 |
| 3C | 8 |
| 4A | 9 |
| 4B | 10 |

**Table 5.** Hyperparameter ranges used in the cross-validation model optimisation.

| Algorithm | Hyperparameter | Range |
| --- | --- | --- |
| Elastic net | Penalty coefficient $\alpha$ | $10^{-8} - 10$ (200 log steps) |
| Elastic net | L1 ratio | 0.1–1.0 (steps of 0.1) |
| SVR | Kernel coefficient $\gamma$ | $10^{-9} - 10^3$ (100 log steps) |
| SVR | Regularisation parameter $C$ | $10^{-3} - 10^3$ (100 log steps) |
| RF | Max. depth | 3 or automatic |
| RF | Num. of estimators | 5, 10, 25, 50, 100 |
| RF | Max. features in split | 0.05, 0.1, 0.2, 0.5, 0.7 |
| RF | Min. samples in split | 2, 3, 6, 10, 12, 15 |

**Table 6.** Optimised hyperparameters used in each of the algorithms and seeds.

| Algorithm | Seed 1 | Seed 2 | Seed 3 | Seed 4 | Seed 5 |  |
| --- | --- | --- | --- | --- | --- | --- |
| Elastic net | Penalty coefficient $\alpha$ | 0.049 | 0.542 | 0.667 | 0.667 | 0.667 |
| Elastic net | L1 ratio | 0.200 | 0.400 | 0.300 | 0.200 | 0.300 |
| SVR | Regularisation parameter $C$ | 869.749 | 2.154 | 10.000 | 869.749 | 657.933 |
| SVR | Kernel coefficient $\gamma$ | 0.011 | 0.033 | 0.011 | 0.011 | 0.000 |
| RF | Max. depth | Automatic | Automatic | 3.0 | 3.0 | 3.0 |
| RF | Max. features in split | 0.7 | 0.5 | 0.1 | 0.1 | 0.1 |
| RF | Min. samples in split | 12.0 | 2.0 | 10.0 | 6.0 | 10.0 |
| RF | Num. estimators | 10.0 | 50.0 | 5.0 | 5.0 | 10.0 |

**Table 7.**
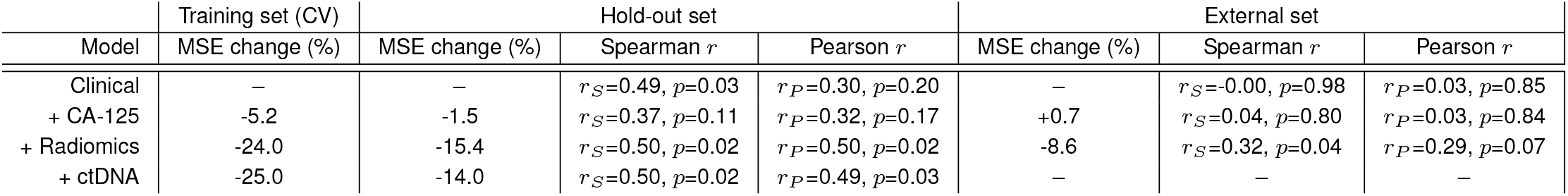
Performance results for the cross-validation training set and the two validation sets.

**Table 8.** List of all non-imaging features used in the predictive models, and their means, medians, standard deviations and ranges for the training, hold-out validation and external validation sets.

| Feature | Training set |  |  |  | Hold-out validation set |  |  |  | External validation set |  |  |  |
| --- | --- | --- | --- | --- | --- | --- | --- | --- | --- | --- | --- | --- |
| | Mean | Median | $\sigma$ | Range | Mean | Median | $\sigma$ | Range | Mean | Median | $\sigma$ | Range |
| Stage | 8.65 | 8.0 | 0.84 | 8-10 | 8.25 | 8.0 | 0.77 | 7-10 | 8.26 | 8.0 | 0.44 | 8-9 |
| Age at diagnosis | 64.43 | 66.5 | 12.19 | 29.0-90.0 | 63.85 | 66.0 | 10.3 | 47.0-83.0 | 63.14 | 65.0 | 12.19 | 35-85 |
| Baseline CA-125 | 2.1e+03 | 972.0 | 2.6e+03 | 11-1.2e+04 | 729.15 | 523.0 | 637.69 | 33-2.1e+03 | 2.3e+03 | 899.5 | 5.1e+03 | 74-3.1e+04 |
| ctDNA tMAD | 0.044 | 0.012 | 0.073 | 0.0047-0.36 | 0.014 | 0.0087 | 0.013 | 0.005-0.057 | — | — | — | — |
| ctDNA TP53 MAF | 0.071 | 0.012 | 0.13 | 0.0-0.64 | 0.014 | 0.0 | 0.025 | 0.0-0.1 | — | — | — | — |
| ctDNA TP53 mutation status | 0.57 | 1.0 | 0.5 | 0.0-1.0 | 0.4 | 0.0 | 0.49 | 0.0-1.0 | — | — | — | — |
| Num. cycles before 2nd CT | 4.056 | 3.0 | 1.95 | 2.0-9.0 | 4.35 | 3.0 | 2.17 | 3.0-9.0 | 3.4 | 3.0 | 0.73 | 3-6 |
| Num. days between 1st CT and treatment start | 25.0 | 26.0 | 13.2 | 1.0-68.0 | 21.55 | 23.5 | 15.61 | 1.0-70.0 | 33.048 | 27.0 | 21.94 | 0.0-99.0 |
| Num. days between 1st and 2nd CTs | 80.97 | 80.5 | 18.59 | 48.0-156.0 | 76.15 | 75.5 | 19.27 | 48.0-143.0 | 84.38 | 78.0 | 27.45 | 45.0-173.0 |
| Received Doxorubicin | 0.042 | 0.0 | 0.2 | 0.0-1.0 | 0.05 | 0.0 | 0.22 | 0.0-1.0 | 0.0 | 0.0 | 0.0 | 0.0-0.0 |
| Paclit. and Carbop. weekly | 0.028 | 0.0 | 0.16 | 0.0-1.0 | 0.0 | 0.0 | 0.0 | 0.0-0.0 | 0.0 | 0.0 | 0.0 | 0.0-0.0 |
| Paclit. and Carbop. 3-weekly | 0.46 | 0.0 | 0.5 | 0.0-1.0 | 0.45 | 0.0 | 0.5 | 0.0-1.0 | 0.93 | 1.0 | 0.26 | 0.0-1.0 |
| Paclit. weekly, Carboplat. 3-weekly | 0.18 | 0.0 | 0.38 | 0.0-1.0 | 0.25 | 0.0 | 0.43 | 0.0-1.0 | 0.0 | 0.0 | 0.0 | 0.0-0.0 |
| Only Carboplatin | 0.24 | 0.0 | 0.42 | 0.0-1.0 | 0.15 | 0.0 | 0.36 | 0.0-1.0 | 0.071 | 0.0 | 0.26 | 0.0-1.0 |
| Received Paclitaxel | 0.72 | 1.0 | 0.45 | 0.0-1.0 | 0.8 | 1.0 | 0.4 | 0.0-1.0 | 0.93 | 1.0 | 0.26 | 0.0-1.0 |

**Table 9.**
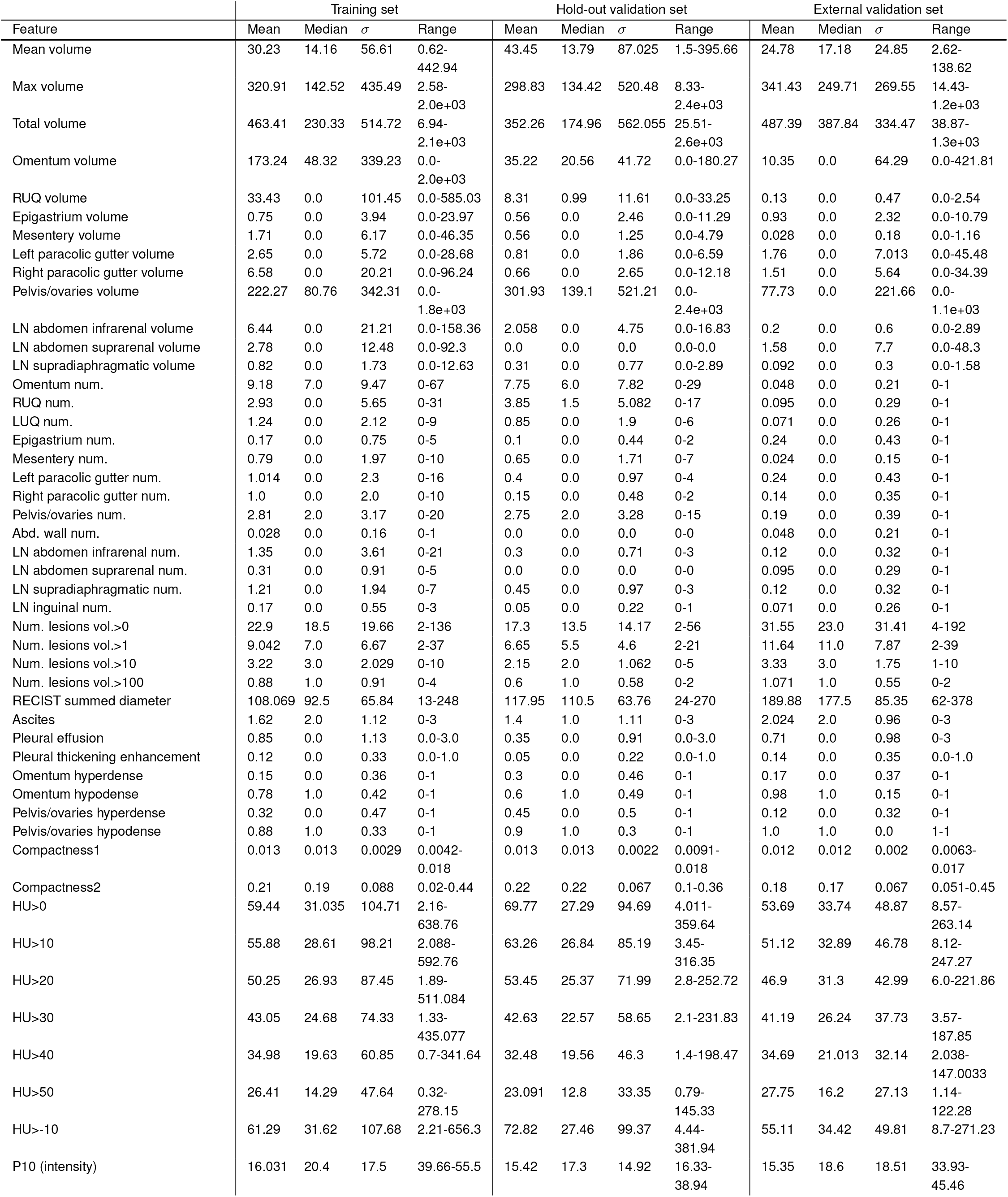
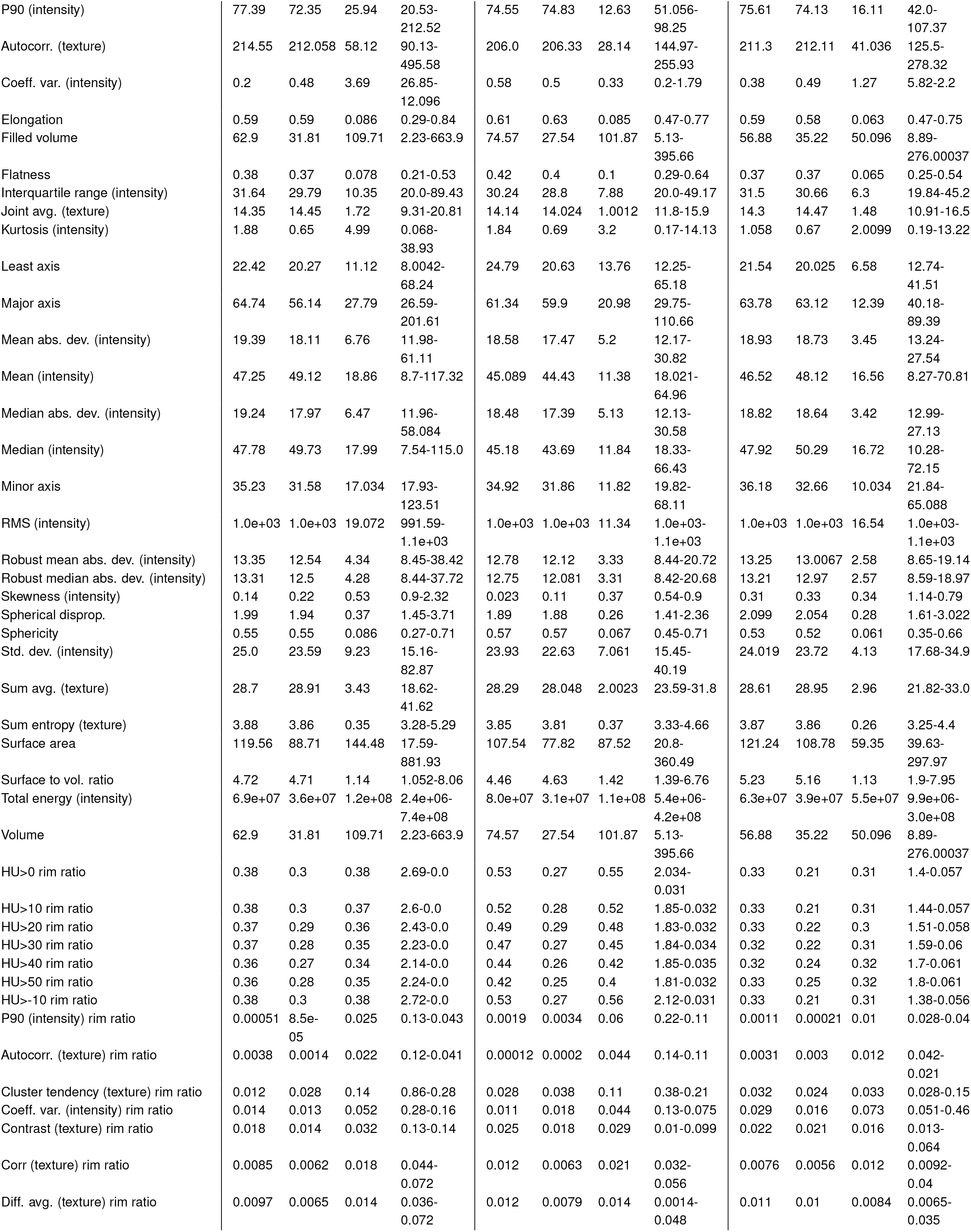
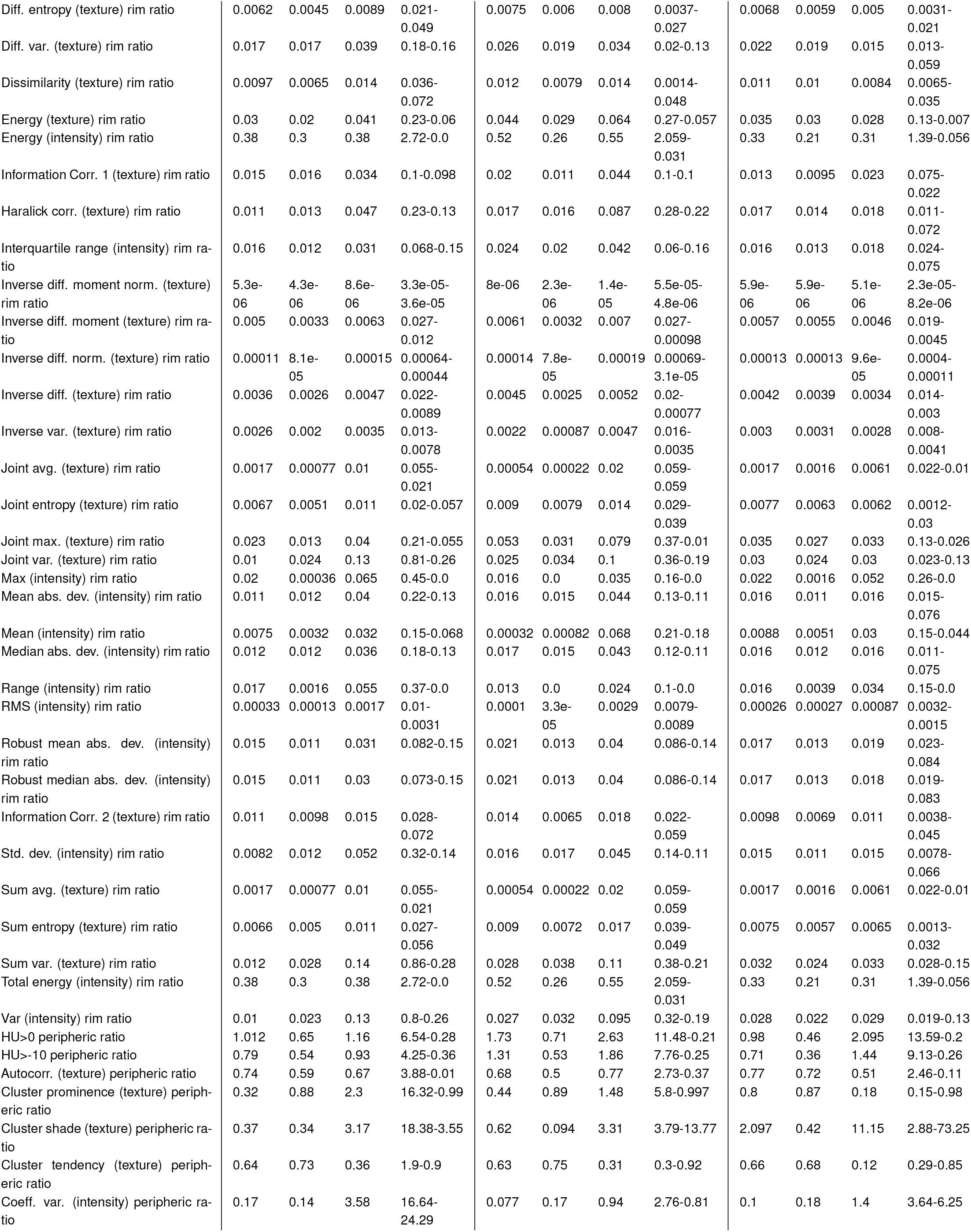
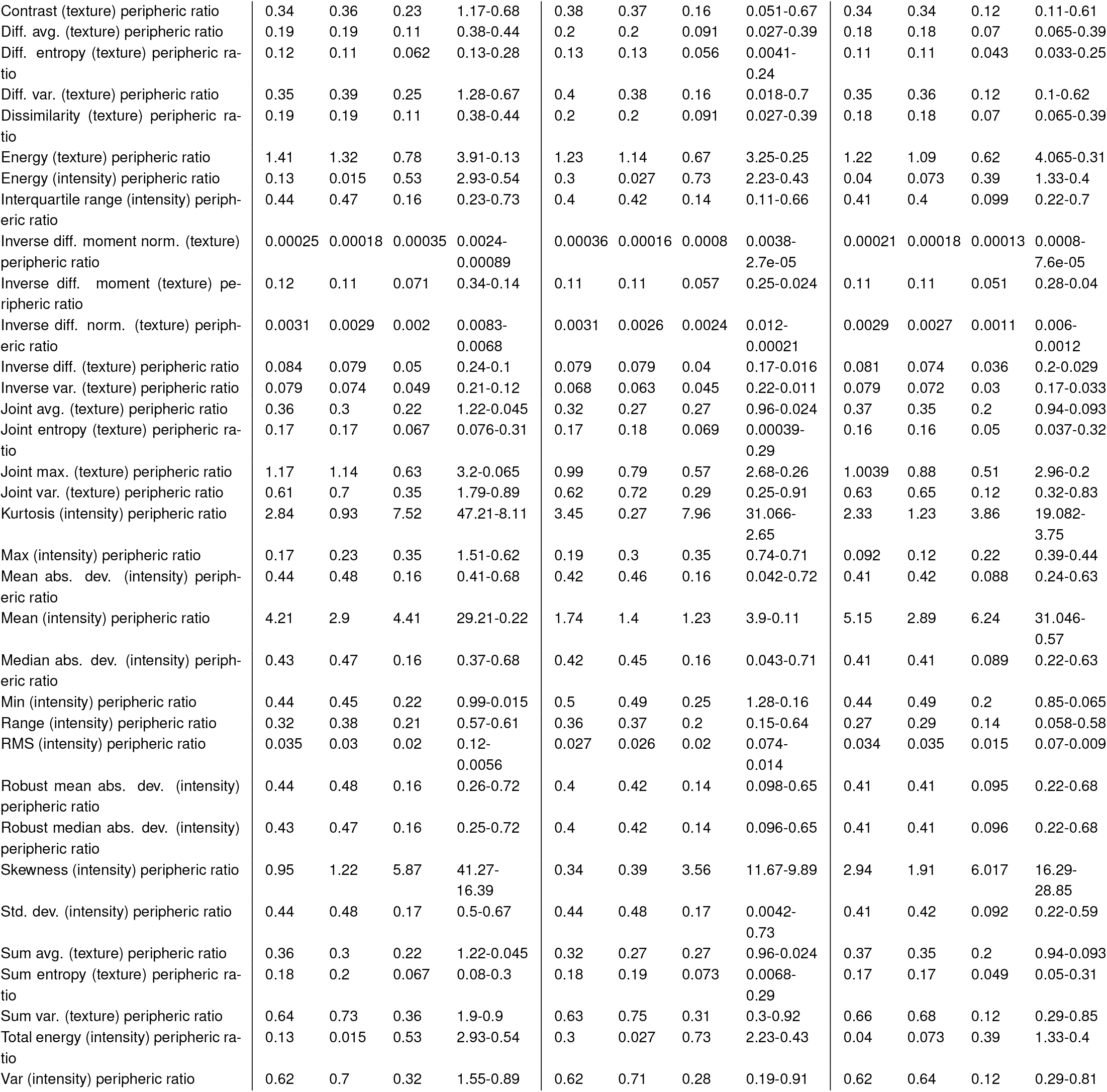
List of all imaging features used in the predictive models, and their means, medians, standard deviations and ranges for the training, hold-out validation and external validation sets.

| Feature | Training set |  |  |  | Hold-out validation set |  |  |  | External validation set |  |  |  |
| --- | --- | --- | --- | --- | --- | --- | --- | --- | --- | --- | --- | --- |
| | Mean | Median | $\sigma$ | Range | Mean | Median | $\sigma$ | Range | Mean | Median | $\sigma$ | Range |
| Mean volume | 30.23 | 14.16 | 56.61 | 0.62-442.94 | 43.45 | 13.79 | 87.025 | 1.5-395.66 | 24.78 | 17.18 | 24.85 | 2.62-138.62 |
| Max volume | 320.91 | 142.52 | 435.49 | 2.58-2.0e+03 | 298.83 | 134.42 | 520.48 | 8.33-2.4e+03 | 341.43 | 249.71 | 269.55 | 14.43-1.2e+03 |
| Total volume | 463.41 | 230.33 | 514.72 | 6.94-2.1e+03 | 352.26 | 174.96 | 562.055 | 25.51-2.6e+03 | 487.39 | 387.84 | 334.47 | 38.87-1.3e+03 |
| Omentum volume | 173.24 | 48.32 | 339.23 | 0.0-2.0e+03 | 35.22 | 20.56 | 41.72 | 0.0-180.27 | 10.35 | 0.0 | 64.29 | 0.0-421.81 |
| RUQ volume | 33.43 | 0.0 | 101.45 | 0.0-585.03 | 8.31 | 0.99 | 11.61 | 0.0-33.25 | 0.13 | 0.0 | 0.47 | 0.0-2.54 |
| Epigastrium volume | 0.75 | 0.0 | 3.94 | 0.0-23.97 | 0.56 | 0.0 | 2.46 | 0.0-11.29 | 0.93 | 0.0 | 2.32 | 0.0-10.79 |
| Mesentery volume | 1.71 | 0.0 | 6.17 | 0.0-46.35 | 0.56 | 0.0 | 1.25 | 0.0-4.79 | 0.028 | 0.0 | 0.18 | 0.0-1.16 |
| Left paracolic gutter volume | 2.65 | 0.0 | 5.72 | 0.0-28.68 | 0.81 | 0.0 | 1.86 | 0.0-6.59 | 1.76 | 0.0 | 7.013 | 0.0-45.48 |
| Right paracolic gutter volume | 6.58 | 0.0 | 20.21 | 0.0-96.24 | 0.66 | 0.0 | 2.65 | 0.0-12.18 | 1.51 | 0.0 | 5.64 | 0.0-34.39 |
| Pelvis/ovaries volume | 222.27 | 80.76 | 342.31 | 0.0-1.8e+03 | 301.93 | 139.1 | 521.21 | 0.0-2.4e+03 | 77.73 | 0.0 | 221.66 | 0.0-1.1e+03 |
| LN abdomen infrarenal volume | 6.44 | 0.0 | 21.21 | 0.0-158.36 | 2.058 | 0.0 | 4.75 | 0.0-16.83 | 0.2 | 0.0 | 0.6 | 0.0-2.89 |
| LN abdomen suprarenal volume | 2.78 | 0.0 | 12.48 | 0.0-92.3 | 0.0 | 0.0 | 0.0 | 0.0-0.0 | 1.58 | 0.0 | 7.7 | 0.0-48.3 |
| LN supradiaphragmatic volume | 0.82 | 0.0 | 1.73 | 0.0-12.63 | 0.31 | 0.0 | 0.77 | 0.0-2.89 | 0.092 | 0.0 | 0.3 | 0.0-1.58 |
| Omentum num. | 9.18 | 7.0 | 9.47 | 0-67 | 7.75 | 6.0 | 7.82 | 0-29 | 0.048 | 0.0 | 0.21 | 0-1 |
| RUQ num. | 2.93 | 0.0 | 5.65 | 0-31 | 3.85 | 1.5 | 5.082 | 0-17 | 0.095 | 0.0 | 0.29 | 0-1 |
| LUQ num. | 1.24 | 0.0 | 2.12 | 0-9 | 0.85 | 0.0 | 1.9 | 0-6 | 0.071 | 0.0 | 0.26 | 0-1 |
| Epigastrium num. | 0.17 | 0.0 | 0.75 | 0-5 | 0.1 | 0.0 | 0.44 | 0-2 | 0.24 | 0.0 | 0.43 | 0-1 |
| Mesentery num. | 0.79 | 0.0 | 1.97 | 0-10 | 0.65 | 0.0 | 1.71 | 0-7 | 0.024 | 0.0 | 0.15 | 0-1 |
| Left paracolic gutter num. | 1.014 | 0.0 | 2.3 | 0-16 | 0.4 | 0.0 | 0.97 | 0-4 | 0.24 | 0.0 | 0.43 | 0-1 |
| Right paracolic gutter num. | 1.0 | 0.0 | 2.0 | 0-10 | 0.15 | 0.0 | 0.48 | 0-2 | 0.14 | 0.0 | 0.35 | 0-1 |
| Pelvis/ovaries num. | 2.81 | 2.0 | 3.17 | 0-20 | 2.75 | 2.0 | 3.28 | 0-15 | 0.19 | 0.0 | 0.39 | 0-1 |
| Abd. wall num. | 0.028 | 0.0 | 0.16 | 0-1 | 0.0 | 0.0 | 0.0 | 0-0 | 0.048 | 0.0 | 0.21 | 0-1 |
| LN abdomen infrarenal num. | 1.35 | 0.0 | 3.61 | 0-21 | 0.3 | 0.0 | 0.71 | 0-3 | 0.12 | 0.0 | 0.32 | 0-1 |
| LN abdomen suprarenal num. | 0.31 | 0.0 | 0.91 | 0-5 | 0.0 | 0.0 | 0.0 | 0-0 | 0.095 | 0.0 | 0.29 | 0-1 |
| LN supradiaphragmatic num. | 1.21 | 0.0 | 1.94 | 0-7 | 0.45 | 0.0 | 0.97 | 0-3 | 0.12 | 0.0 | 0.32 | 0-1 |
| LN inguinal num. | 0.17 | 0.0 | 0.55 | 0-3 | 0.05 | 0.0 | 0.22 | 0-1 | 0.071 | 0.0 | 0.26 | 0-1 |
| Num. lesions vol.>0 | 22.9 | 18.5 | 19.66 | 2-136 | 17.3 | 13.5 | 14.17 | 2-56 | 31.55 | 23.0 | 31.41 | 4-192 |
| Num. lesions vol.>1 | 9.042 | 7.0 | 6.67 | 2-37 | 6.65 | 5.5 | 4.6 | 2-21 | 11.64 | 11.0 | 7.87 | 2-39 |
| Num. lesions vol.>10 | 3.22 | 3.0 | 2.029 | 0-10 | 2.15 | 2.0 | 1.062 | 0-5 | 3.33 | 3.0 | 1.75 | 1-10 |
| Num. lesions vol.>100 | 0.88 | 1.0 | 0.91 | 0-4 | 0.6 | 1.0 | 0.58 | 0-2 | 1.071 | 1.0 | 0.55 | 0-2 |
| RECIST summed diameter | 108.069 | 92.5 | 65.84 | 13-248 | 117.95 | 110.5 | 63.76 | 24-270 | 189.88 | 177.5 | 85.35 | 62-378 |
| Ascites | 1.62 | 2.0 | 1.12 | 0-3 | 1.4 | 1.0 | 1.11 | 0-3 | 2.024 | 2.0 | 0.96 | 0-3 |
| Pleural effusion | 0.85 | 0.0 | 1.13 | 0.0-3.0 | 0.35 | 0.0 | 0.91 | 0.0-3.0 | 0.71 | 0.0 | 0.98 | 0-3 |
| Pleural thickening enhancement | 0.12 | 0.0 | 0.33 | 0.0-1.0 | 0.05 | 0.0 | 0.22 | 0.0-1.0 | 0.14 | 0.0 | 0.35 | 0.0-1.0 |
| Omentum hyperdense | 0.15 | 0.0 | 0.36 | 0-1 | 0.3 | 0.0 | 0.46 | 0-1 | 0.17 | 0.0 | 0.37 | 0-1 |
| Omentum hypodense | 0.78 | 1.0 | 0.42 | 0-1 | 0.6 | 1.0 | 0.49 | 0-1 | 0.98 | 1.0 | 0.15 | 0-1 |
| Pelvis/ovaries hyperdense | 0.32 | 0.0 | 0.47 | 0-1 | 0.45 | 0.0 | 0.5 | 0-1 | 0.12 | 0.0 | 0.32 | 0-1 |
| Pelvis/ovaries hypodense | 0.88 | 1.0 | 0.33 | 0-1 | 0.9 | 1.0 | 0.3 | 0-1 | 1.0 | 1.0 | 0.0 | 1-1 |
| Compactness1 | 0.013 | 0.013 | 0.0029 | 0.0042-0.018 | 0.013 | 0.013 | 0.0022 | 0.0091-0.018 | 0.012 | 0.012 | 0.002 | 0.0063-0.017 |
| Compactness2 | 0.21 | 0.19 | 0.088 | 0.02-0.44 | 0.22 | 0.22 | 0.067 | 0.1-0.36 | 0.18 | 0.17 | 0.067 | 0.051-0.45 |
| HU>0 | 59.44 | 31.035 | 104.71 | 2.16-638.76 | 69.77 | 27.29 | 94.69 | 4.011-359.64 | 53.69 | 33.74 | 48.87 | 8.57-263.14 |
| HU>10 | 55.88 | 28.61 | 98.21 | 2.088-592.76 | 63.26 | 26.84 | 85.19 | 3.45-316.35 | 51.12 | 32.89 | 46.78 | 8.12-247.27 |
| HU>20 | 50.25 | 26.93 | 87.45 | 1.89-511.084 | 53.45 | 25.37 | 71.99 | 2.8-252.72 | 46.9 | 31.3 | 42.99 | 6.0-221.86 |
| HU>30 | 43.05 | 24.68 | 74.33 | 1.33-435.077 | 42.63 | 22.57 | 58.65 | 2.1-231.83 | 41.19 | 26.24 | 37.73 | 3.57-187.85 |
| HU>40 | 34.98 | 19.63 | 60.85 | 0.7-341.64 | 32.48 | 19.56 | 46.3 | 1.4-198.47 | 34.69 | 21.013 | 32.14 | 2.038-147.0033 |
| HU>50 | 26.41 | 14.29 | 47.64 | 0.32-278.15 | 23.091 | 12.8 | 33.35 | 0.79-145.33 | 27.75 | 16.2 | 27.13 | 1.14-122.28 |
| HU>-10 | 61.29 | 31.62 | 107.68 | 2.21-656.3 | 72.82 | 27.46 | 99.37 | 4.44-381.94 | 55.11 | 34.42 | 49.81 | 8.7-271.23 |
| P10 (intensity) | 16.031 | 20.4 | 17.5 | 39.66-55.5 | 15.42 | 17.3 | 14.92 | 16.33-38.94 | 15.35 | 18.6 | 18.51 | 33.93-45.46 |
| P90 (intensity) | 77.39 | 72.35 | 25.94 | 20.53-212.52 | 74.55 | 74.83 | 12.63 | 51.056-98.25 | 75.61 | 74.13 | 16.11 | 42.0-107.37 |
| Autocorr. (texture) | 214.55 | 212.058 | 58.12 | 90.13-495.58 | 206.0 | 206.33 | 28.14 | 144.97-255.93 | 211.3 | 212.11 | 41.036 | 125.5-278.32 |
| Coeff. var. (intensity) | 0.2 | 0.48 | 3.69 | 26.85-12.096 | 0.58 | 0.5 | 0.33 | 0.2-1.79 | 0.38 | 0.49 | 1.27 | 5.82-2.2 |
| Elongation | 0.59 | 0.59 | 0.086 | 0.29-0.84 | 0.61 | 0.63 | 0.085 | 0.47-0.77 | 0.59 | 0.58 | 0.063 | 0.47-0.75 |
| Filled volume | 62.9 | 31.81 | 109.71 | 2.23-663.9 | 74.57 | 27.54 | 101.87 | 5.13-395.66 | 56.88 | 35.22 | 50.096 | 8.89-276.00037 |
| Flatness | 0.38 | 0.37 | 0.078 | 0.21-0.53 | 0.42 | 0.4 | 0.1 | 0.29-0.64 | 0.37 | 0.37 | 0.065 | 0.25-0.54 |
| Interquartile range (intensity) | 31.64 | 29.79 | 10.35 | 20.0-89.43 | 30.24 | 28.8 | 7.88 | 20.0-49.17 | 31.5 | 30.66 | 6.3 | 19.84-45.2 |
| Joint avg. (texture) | 14.35 | 14.45 | 1.72 | 9.31-20.81 | 14.14 | 14.024 | 1.0012 | 11.8-15.9 | 14.3 | 14.47 | 1.48 | 10.91-16.5 |
| Kurtosis (intensity) | 1.88 | 0.65 | 4.99 | 0.068-38.93 | 1.84 | 0.69 | 3.2 | 0.17-14.13 | 1.058 | 0.67 | 2.0099 | 0.19-13.22 |
| Least axis | 22.42 | 20.27 | 11.12 | 8.0042-68.24 | 24.79 | 20.63 | 13.76 | 12.25-65.18 | 21.54 | 20.025 | 6.58 | 12.74-41.51 |
| Major axis | 64.74 | 56.14 | 27.79 | 26.59-201.61 | 61.34 | 59.9 | 20.98 | 29.75-110.66 | 63.78 | 63.12 | 12.39 | 40.18-89.39 |
| Mean abs. dev. (intensity) | 19.39 | 18.11 | 6.76 | 11.98-61.11 | 18.58 | 17.47 | 5.2 | 12.17-30.82 | 18.93 | 18.73 | 3.45 | 13.24-27.54 |
| Mean (intensity) | 47.25 | 49.12 | 18.86 | 8.7-117.32 | 45.089 | 44.43 | 11.38 | 18.021-64.96 | 46.52 | 48.12 | 16.56 | 8.27-70.81 |
| Median abs. dev. (intensity) | 19.24 | 17.97 | 6.47 | 11.96-58.084 | 18.48 | 17.39 | 5.13 | 12.13-30.58 | 18.82 | 18.64 | 3.42 | 12.99-27.13 |
| Median (intensity) | 47.78 | 49.73 | 17.99 | 7.54-115.0 | 45.18 | 43.69 | 11.84 | 18.33-66.43 | 47.92 | 50.29 | 16.72 | 10.28-72.15 |
| Minor axis | 35.23 | 31.58 | 17.034 | 17.93-123.51 | 34.92 | 31.86 | 11.82 | 19.82-68.11 | 36.18 | 32.66 | 10.034 | 21.84-65.088 |
| RMS (intensity) | 1.0e+03 | 1.0e+03 | 19.072 | 991.59-1.1e+03 | 1.0e+03 | 1.0e+03 | 11.34 | 1.0e+03-1.1e+03 | 1.0e+03 | 1.0e+03 | 16.54 | 1.0e+03-1.1e+03 |
| Robust mean abs. dev. (intensity) | 13.35 | 12.54 | 4.34 | 8.45-38.42 | 12.78 | 12.12 | 3.33 | 8.44-20.72 | 13.25 | 13.0067 | 2.58 | 8.65-19.14 |
| Robust median abs. dev. (intensity) | 13.31 | 12.5 | 4.28 | 8.44-37.72 | 12.75 | 12.081 | 3.31 | 8.42-20.68 | 13.21 | 12.97 | 2.57 | 8.59-18.97 |
| Skewness (intensity) | 0.14 | 0.22 | 0.53 | 0.9-2.32 | 0.023 | 0.11 | 0.37 | 0.54-0.9 | 0.31 | 0.33 | 0.34 | 1.14-0.79 |
| Spherical disprop. | 1.99 | 1.94 | 0.37 | 1.45-3.71 | 1.89 | 1.88 | 0.26 | 1.41-2.36 | 2.099 | 2.054 | 0.28 | 1.61-3.022 |
| Sphericity | 0.55 | 0.55 | 0.086 | 0.27-0.71 | 0.57 | 0.57 | 0.067 | 0.45-0.71 | 0.53 | 0.52 | 0.061 | 0.35-0.66 |
| Std. dev. (intensity) | 25.0 | 23.59 | 9.23 | 15.16-82.87 | 23.93 | 22.63 | 7.061 | 15.45-40.19 | 24.019 | 23.72 | 4.13 | 17.68-34.9 |
| Sum avg. (texture) | 28.7 | 28.91 | 3.43 | 18.62-41.62 | 28.29 | 28.048 | 2.0023 | 23.59-31.8 | 28.61 | 28.95 | 2.96 | 21.82-33.0 |
| Sum entropy (texture) | 3.88 | 3.86 | 0.35 | 3.28-5.29 | 3.85 | 3.81 | 0.37 | 3.33-4.66 | 3.87 | 3.86 | 0.26 | 3.25-4.4 |
| Surface area | 119.56 | 88.71 | 144.48 | 17.59-881.93 | 107.54 | 77.82 | 87.52 | 20.8-360.49 | 121.24 | 108.78 | 59.35 | 39.63-297.97 |
| Surface to vol. ratio | 4.72 | 4.71 | 1.14 | 1.052-8.06 | 4.46 | 4.63 | 1.42 | 1.39-6.76 | 5.23 | 5.16 | 1.13 | 1.9-7.95 |
| Total energy (intensity) | 6.9e+07 | 3.6e+07 | 1.2e+08 | 2.4e+06-7.4e+08 | 8.0e+07 | 3.1e+07 | 1.1e+08 | 5.4e+06-4.2e+08 | 6.3e+07 | 3.9e+07 | 5.5e+07 | 9.9e+06-3.0e+08 |
| Volume | 62.9 | 31.81 | 109.71 | 2.23-663.9 | 74.57 | 27.54 | 101.87 | 5.13-395.66 | 56.88 | 35.22 | 50.096 | 8.89-276.00037 |
| HU>0 rim ratio | 0.38 | 0.3 | 0.38 | 2.69-0.0 | 0.53 | 0.27 | 0.55 | 2.034-0.031 | 0.33 | 0.21 | 0.31 | 1.4-0.057 |
| HU>10 rim ratio | 0.38 | 0.3 | 0.37 | 2.6-0.0 | 0.52 | 0.28 | 0.52 | 1.85-0.032 | 0.33 | 0.21 | 0.31 | 1.44-0.057 |
| HU>20 rim ratio | 0.37 | 0.29 | 0.36 | 2.43-0.0 | 0.49 | 0.29 | 0.48 | 1.83-0.032 | 0.33 | 0.22 | 0.3 | 1.51-0.058 |
| HU>30 rim ratio | 0.37 | 0.28 | 0.35 | 2.23-0.0 | 0.47 | 0.27 | 0.45 | 1.84-0.034 | 0.32 | 0.22 | 0.31 | 1.59-0.06 |
| HU>40 rim ratio | 0.36 | 0.27 | 0.34 | 2.14-0.0 | 0.44 | 0.26 | 0.42 | 1.85-0.035 | 0.32 | 0.24 | 0.32 | 1.7-0.061 |
| HU>50 rim ratio | 0.36 | 0.28 | 0.35 | 2.24-0.0 | 0.42 | 0.25 | 0.4 | 1.81-0.032 | 0.33 | 0.25 | 0.32 | 1.8-0.061 |
| HU>-10 rim ratio | 0.38 | 0.3 | 0.38 | 2.72-0.0 | 0.53 | 0.27 | 0.56 | 2.12-0.031 | 0.33 | 0.21 | 0.31 | 1.38-0.056 |
| P90 (intensity) rim ratio | 0.00051 | 8.5e-05 | 0.025 | 0.13-0.043 | 0.0019 | 0.0034 | 0.06 | 0.22-0.11 | 0.0011 | 0.00021 | 0.01 | 0.028-0.04 |
| Autocorr. (texture) rim ratio | 0.0038 | 0.0014 | 0.022 | 0.12-0.041 | 0.00012 | 0.0002 | 0.044 | 0.14-0.11 | 0.0031 | 0.003 | 0.012 | 0.042-0.021 |
| Cluster tendency (texture) rim ratio | 0.012 | 0.028 | 0.14 | 0.86-0.28 | 0.028 | 0.038 | 0.11 | 0.38-0.21 | 0.032 | 0.024 | 0.033 | 0.028-0.15 |
| Coeff. var. (intensity) rim ratio | 0.014 | 0.013 | 0.052 | 0.28-0.16 | 0.011 | 0.018 | 0.044 | 0.13-0.075 | 0.029 | 0.016 | 0.073 | 0.051-0.46 |
| Contrast (texture) rim ratio | 0.018 | 0.014 | 0.032 | 0.13-0.14 | 0.025 | 0.018 | 0.029 | 0.01-0.099 | 0.022 | 0.021 | 0.016 | 0.013-0.064 |
| Corr (texture) rim ratio | 0.0085 | 0.0062 | 0.018 | 0.044-0.072 | 0.012 | 0.0063 | 0.021 | 0.032-0.056 | 0.0076 | 0.0056 | 0.012 | 0.0092-0.04 |
| Diff. avg. (texture) rim ratio | 0.0097 | 0.0065 | 0.014 | 0.036-0.072 | 0.012 | 0.0079 | 0.014 | 0.0014-0.048 | 0.011 | 0.01 | 0.0084 | 0.0065-0.035 |
| Diff. entropy (texture) rim ratio | 0.0062 | 0.0045 | 0.0089 | 0.021-0.049 | 0.0075 | 0.006 | 0.008 | 0.0037-0.027 | 0.0068 | 0.0059 | 0.005 | 0.0031-0.021 |
| Diff. var. (texture) rim ratio | 0.017 | 0.017 | 0.039 | 0.18-0.16 | 0.026 | 0.019 | 0.034 | 0.02-0.13 | 0.022 | 0.019 | 0.015 | 0.013-0.059 |
| Dissimilarity (texture) rim ratio | 0.0097 | 0.0065 | 0.014 | 0.036-0.072 | 0.012 | 0.0079 | 0.014 | 0.0014-0.048 | 0.011 | 0.01 | 0.0084 | 0.0065-0.035 |
| Energy (texture) rim ratio | 0.03 | 0.02 | 0.041 | 0.23-0.06 | 0.044 | 0.029 | 0.064 | 0.27-0.057 | 0.035 | 0.03 | 0.028 | 0.13-0.007 |
| Energy (intensity) rim ratio | 0.38 | 0.3 | 0.38 | 2.72-0.0 | 0.52 | 0.26 | 0.55 | 2.059-0.031 | 0.33 | 0.21 | 0.31 | 1.39-0.056 |
| Information Corr. 1 (texture) rim ratio | 0.015 | 0.016 | 0.034 | 0.1-0.098 | 0.02 | 0.011 | 0.044 | 0.1-0.1 | 0.013 | 0.0095 | 0.023 | 0.075-0.022 |
| Haralick corr. (texture) rim ratio | 0.011 | 0.013 | 0.047 | 0.23-0.13 | 0.017 | 0.016 | 0.087 | 0.28-0.22 | 0.017 | 0.014 | 0.018 | 0.011-0.072 |
| Interquartile range (intensity) rim ratio | 0.016 | 0.012 | 0.031 | 0.068-0.15 | 0.024 | 0.02 | 0.042 | 0.06-0.16 | 0.016 | 0.013 | 0.018 | 0.024-0.075 |
| Inverse diff. moment norm. (texture) rim ratio | 5.3e-06 | 4.3e-06 | 8.6e-06 | 3.3e-05-3.6e-05 | 8e-06 | 2.3e-06 | 1.4e-05 | 5.5e-05-4.8e-06 | 5.9e-06 | 5.9e-06 | 5.1e-06 | 2.3e-05-8.2e-06 |
| Inverse diff. moment (texture) rim ratio | 0.005 | 0.0033 | 0.0063 | 0.027-0.012 | 0.0061 | 0.0032 | 0.007 | 0.027-0.00098 | 0.0057 | 0.0055 | 0.0046 | 0.019-0.0045 |
| Inverse diff. norm. (texture) rim ratio | 0.00011 | 8.1e-05 | 0.00015 | 0.00064-0.00044 | 0.00014 | 7.8e-05 | 0.00019 | 0.00069-3.1e-05 | 0.00013 | 0.00013 | 9.6e-05 | 0.0004-0.00011 |
| Inverse diff. (texture) rim ratio | 0.0036 | 0.0026 | 0.0047 | 0.022-0.0089 | 0.0045 | 0.0025 | 0.0052 | 0.02-0.00077 | 0.0042 | 0.0039 | 0.0034 | 0.014-0.003 |
| Inverse var. (texture) rim ratio | 0.0026 | 0.002 | 0.0035 | 0.013-0.0078 | 0.0022 | 0.00087 | 0.0047 | 0.016-0.0035 | 0.003 | 0.0031 | 0.0028 | 0.008-0.0041 |
| Joint avg. (texture) rim ratio | 0.0017 | 0.00077 | 0.01 | 0.055-0.021 | 0.00054 | 0.00022 | 0.02 | 0.059-0.059 | 0.0017 | 0.0016 | 0.0061 | 0.022-0.01 |
| Joint entropy (texture) rim ratio | 0.0067 | 0.0051 | 0.011 | 0.02-0.057 | 0.009 | 0.0079 | 0.014 | 0.029-0.039 | 0.0077 | 0.0063 | 0.0062 | 0.0012-0.03 |
| Joint max. (texture) rim ratio | 0.023 | 0.013 | 0.04 | 0.21-0.055 | 0.053 | 0.031 | 0.079 | 0.37-0.01 | 0.035 | 0.027 | 0.033 | 0.13-0.026 |
| Joint var. (texture) rim ratio | 0.01 | 0.024 | 0.13 | 0.81-0.26 | 0.025 | 0.034 | 0.1 | 0.36-0.19 | 0.03 | 0.024 | 0.03 | 0.023-0.13 |
| Max (intensity) rim ratio | 0.02 | 0.00036 | 0.065 | 0.45-0.0 | 0.016 | 0.0 | 0.035 | 0.16-0.0 | 0.022 | 0.0016 | 0.052 | 0.26-0.0 |
| Mean abs. dev. (intensity) rim ratio | 0.011 | 0.012 | 0.04 | 0.22-0.13 | 0.016 | 0.015 | 0.044 | 0.13-0.11 | 0.016 | 0.011 | 0.016 | 0.015-0.076 |
| Mean (intensity) rim ratio | 0.0075 | 0.0032 | 0.032 | 0.15-0.068 | 0.00032 | 0.00082 | 0.068 | 0.21-0.18 | 0.0088 | 0.0051 | 0.03 | 0.15-0.044 |
| Median abs. dev. (intensity) rim ratio | 0.012 | 0.012 | 0.036 | 0.18-0.13 | 0.017 | 0.015 | 0.043 | 0.12-0.11 | 0.016 | 0.012 | 0.016 | 0.011-0.075 |
| Range (intensity) rim ratio | 0.017 | 0.0016 | 0.055 | 0.37-0.0 | 0.013 | 0.0 | 0.024 | 0.1-0.0 | 0.016 | 0.0039 | 0.034 | 0.15-0.0 |
| RMS (intensity) rim ratio | 0.00033 | 0.00013 | 0.0017 | 0.01-0.0031 | 0.0001 | 3.3e-05 | 0.0029 | 0.0079-0.0089 | 0.00026 | 0.00027 | 0.00087 | 0.0032-0.0015 |
| Robust mean abs. dev. (intensity) rim ratio | 0.015 | 0.011 | 0.031 | 0.082-0.15 | 0.021 | 0.013 | 0.04 | 0.086-0.14 | 0.017 | 0.013 | 0.019 | 0.023-0.084 |
| Robust median abs. dev. (intensity) rim ratio | 0.015 | 0.011 | 0.03 | 0.073-0.15 | 0.021 | 0.013 | 0.04 | 0.086-0.14 | 0.017 | 0.013 | 0.018 | 0.019-0.083 |
| Information Corr. 2 (texture) rim ratio | 0.011 | 0.0098 | 0.015 | 0.028-0.072 | 0.014 | 0.0065 | 0.018 | 0.022-0.059 | 0.0098 | 0.0069 | 0.011 | 0.0038-0.045 |
| Std. dev. (intensity) rim ratio | 0.0082 | 0.012 | 0.052 | 0.32-0.14 | 0.016 | 0.017 | 0.045 | 0.14-0.11 | 0.015 | 0.011 | 0.015 | 0.0078-0.066 |
| Sum avg. (texture) rim ratio | 0.0017 | 0.00077 | 0.01 | 0.055-0.021 | 0.00054 | 0.00022 | 0.02 | 0.059-0.059 | 0.0017 | 0.0016 | 0.0061 | 0.022-0.01 |
| Sum entropy (texture) rim ratio | 0.0066 | 0.005 | 0.011 | 0.027-0.056 | 0.009 | 0.0072 | 0.017 | 0.039-0.049 | 0.0075 | 0.0057 | 0.0065 | 0.0013-0.032 |
| Sum var. (texture) rim ratio | 0.012 | 0.028 | 0.14 | 0.86-0.28 | 0.028 | 0.038 | 0.11 | 0.38-0.21 | 0.032 | 0.024 | 0.033 | 0.028-0.15 |
| Total energy (intensity) rim ratio | 0.38 | 0.3 | 0.38 | 2.72-0.0 | 0.52 | 0.26 | 0.55 | 2.059-0.031 | 0.33 | 0.21 | 0.31 | 1.39-0.056 |
| Var (intensity) rim ratio | 0.01 | 0.023 | 0.13 | 0.8-0.26 | 0.027 | 0.032 | 0.095 | 0.32-0.19 | 0.028 | 0.022 | 0.029 | 0.019-0.13 |
| HU>0 peripheric ratio | 1.012 | 0.65 | 1.16 | 6.54-0.28 | 1.73 | 0.71 | 2.63 | 11.48-0.21 | 0.98 | 0.46 | 2.095 | 13.59-0.2 |
| HU>-10 peripheric ratio | 0.79 | 0.54 | 0.93 | 4.25-0.36 | 1.31 | 0.53 | 1.86 | 7.76-0.25 | 0.71 | 0.36 | 1.44 | 9.13-0.26 |
| Autocorr. (texture) peripheric ratio | 0.74 | 0.59 | 0.67 | 3.88-0.01 | 0.68 | 0.5 | 0.77 | 2.73-0.37 | 0.77 | 0.72 | 0.51 | 2.46-0.11 |
| Cluster prominence (texture) peripheric ratio | 0.32 | 0.88 | 2.3 | 16.32-0.99 | 0.44 | 0.89 | 1.48 | 5.8-0.997 | 0.8 | 0.87 | 0.18 | 0.15-0.98 |
| Cluster shade (texture) peripheric ratio | 0.37 | 0.34 | 3.17 | 18.38-3.55 | 0.62 | 0.094 | 3.31 | 3.79-13.77 | 2.097 | 0.42 | 11.15 | 2.88-73.25 |
| Cluster tendency (texture) peripheric ratio | 0.64 | 0.73 | 0.36 | 1.9-0.9 | 0.63 | 0.75 | 0.31 | 0.3-0.92 | 0.66 | 0.68 | 0.12 | 0.29-0.85 |
| Coeff. var. (intensity) peripheric ratio | 0.17 | 0.14 | 3.58 | 16.64-24.29 | 0.077 | 0.17 | 0.94 | 2.76-0.81 | 0.1 | 0.18 | 1.4 | 3.64-6.25 |
| Contrast (texture) peripheric ratio | 0.34 | 0.36 | 0.23 | 1.17-0.68 | 0.38 | 0.37 | 0.16 | 0.051-0.67 | 0.34 | 0.34 | 0.12 | 0.11-0.61 |
| Diff. avg. (texture) peripheric ratio | 0.19 | 0.19 | 0.11 | 0.38-0.44 | 0.2 | 0.2 | 0.091 | 0.027-0.39 | 0.18 | 0.18 | 0.07 | 0.065-0.39 |
| Diff. entropy (texture) peripheric ratio | 0.12 | 0.11 | 0.062 | 0.13-0.28 | 0.13 | 0.13 | 0.056 | 0.0041-0.24 | 0.11 | 0.11 | 0.043 | 0.033-0.25 |
| Diff. var. (texture) peripheric ratio | 0.35 | 0.39 | 0.25 | 1.28-0.67 | 0.4 | 0.38 | 0.16 | 0.018-0.7 | 0.35 | 0.36 | 0.12 | 0.1-0.62 |
| Dissimilarity (texture) peripheric ratio | 0.19 | 0.19 | 0.11 | 0.38-0.44 | 0.2 | 0.2 | 0.091 | 0.027-0.39 | 0.18 | 0.18 | 0.07 | 0.065-0.39 |
| Energy (texture) peripheric ratio | 1.41 | 1.32 | 0.78 | 3.91-0.13 | 1.23 | 1.14 | 0.67 | 3.25-0.25 | 1.22 | 1.09 | 0.62 | 4.065-0.31 |
| Energy (intensity) peripheric ratio | 0.13 | 0.015 | 0.53 | 2.93-0.54 | 0.3 | 0.027 | 0.73 | 2.23-0.43 | 0.04 | 0.073 | 0.39 | 1.33-0.4 |
| Interquartile range (intensity) peripheric ratio | 0.44 | 0.47 | 0.16 | 0.23-0.73 | 0.4 | 0.42 | 0.14 | 0.11-0.66 | 0.41 | 0.4 | 0.099 | 0.22-0.7 |
| Inverse diff. moment norm. (texture) peripheric ratio | 0.00025 | 0.00018 | 0.00035 | 0.0024-0.00089 | 0.00036 | 0.00016 | 0.0008 | 0.0038-2.7e-05 | 0.00021 | 0.00018 | 0.00013 | 0.0008-7.6e-05 |
| Inverse diff. moment (texture) peripheric ratio | 0.12 | 0.11 | 0.071 | 0.34-0.14 | 0.11 | 0.11 | 0.057 | 0.25-0.024 | 0.11 | 0.11 | 0.051 | 0.28-0.04 |
| Inverse diff. norm. (texture) peripheric ratio | 0.0031 | 0.0029 | 0.002 | 0.0083-0.0068 | 0.0031 | 0.0026 | 0.0024 | 0.012-0.00021 | 0.0029 | 0.0027 | 0.0011 | 0.006-0.0012 |
| Inverse diff. (texture) peripheric ratio | 0.084 | 0.079 | 0.05 | 0.24-0.1 | 0.079 | 0.079 | 0.04 | 0.17-0.016 | 0.081 | 0.074 | 0.036 | 0.2-0.029 |
| Inverse var. (texture) peripheric ratio | 0.079 | 0.074 | 0.049 | 0.21-0.12 | 0.068 | 0.063 | 0.045 | 0.22-0.011 | 0.079 | 0.072 | 0.03 | 0.17-0.033 |
| Joint avg. (texture) peripheric ratio | 0.36 | 0.3 | 0.22 | 1.22-0.045 | 0.32 | 0.27 | 0.27 | 0.96-0.024 | 0.37 | 0.35 | 0.2 | 0.94-0.093 |
| Joint entropy (texture) peripheric ratio | 0.17 | 0.17 | 0.067 | 0.076-0.31 | 0.17 | 0.18 | 0.069 | 0.00039-0.29 | 0.16 | 0.16 | 0.05 | 0.037-0.32 |
| Joint max. (texture) peripheric ratio | 1.17 | 1.14 | 0.63 | 3.2-0.065 | 0.99 | 0.79 | 0.57 | 2.68-0.26 | 1.0039 | 0.88 | 0.51 | 2.96-0.2 |
| Joint var. (texture) peripheric ratio | 0.61 | 0.7 | 0.35 | 1.79-0.89 | 0.62 | 0.72 | 0.29 | 0.25-0.91 | 0.63 | 0.65 | 0.12 | 0.32-0.83 |
| Kurtosis (intensity) peripheric ratio | 2.84 | 0.93 | 7.52 | 47.21-8.11 | 3.45 | 0.27 | 7.96 | 31.066-2.65 | 2.33 | 1.23 | 3.86 | 19.082-3.75 |
| Max (intensity) peripheric ratio | 0.17 | 0.23 | 0.35 | 1.51-0.62 | 0.19 | 0.3 | 0.35 | 0.74-0.71 | 0.092 | 0.12 | 0.22 | 0.39-0.44 |
| Mean abs. dev. (intensity) peripheric ratio | 0.44 | 0.48 | 0.16 | 0.41-0.68 | 0.42 | 0.46 | 0.16 | 0.042-0.72 | 0.41 | 0.42 | 0.088 | 0.24-0.63 |
| Mean (intensity) peripheric ratio | 4.21 | 2.9 | 4.41 | 29.21-0.22 | 1.74 | 1.4 | 1.23 | 3.9-0.11 | 5.15 | 2.89 | 6.24 | 31.046-0.57 |
| Median abs. dev. (intensity) peripheric ratio | 0.43 | 0.47 | 0.16 | 0.37-0.68 | 0.42 | 0.45 | 0.16 | 0.043-0.71 | 0.41 | 0.41 | 0.089 | 0.22-0.63 |
| Min (intensity) peripheric ratio | 0.44 | 0.45 | 0.22 | 0.99-0.015 | 0.5 | 0.49 | 0.25 | 1.28-0.16 | 0.44 | 0.49 | 0.2 | 0.85-0.065 |
| Range (intensity) peripheric ratio | 0.32 | 0.38 | 0.21 | 0.57-0.61 | 0.36 | 0.37 | 0.2 | 0.15-0.64 | 0.27 | 0.29 | 0.14 | 0.058-0.58 |
| RMS (intensity) peripheric ratio | 0.035 | 0.03 | 0.02 | 0.12-0.0056 | 0.027 | 0.026 | 0.02 | 0.074-0.014 | 0.034 | 0.035 | 0.015 | 0.07-0.009 |
| Robust mean abs. dev. (intensity) peripheric ratio | 0.44 | 0.48 | 0.16 | 0.26-0.72 | 0.4 | 0.42 | 0.14 | 0.098-0.65 | 0.41 | 0.41 | 0.095 | 0.22-0.68 |
| Robust median abs. dev. (intensity) peripheric ratio | 0.43 | 0.47 | 0.16 | 0.25-0.72 | 0.4 | 0.42 | 0.14 | 0.096-0.65 | 0.41 | 0.41 | 0.096 | 0.22-0.68 |
| Skewness (intensity) peripheric ratio | 0.95 | 1.22 | 5.87 | 41.27-16.39 | 0.34 | 0.39 | 3.56 | 11.67-9.89 | 2.94 | 1.91 | 6.017 | 16.29-28.85 |
| Std. dev. (intensity) peripheric ratio | 0.44 | 0.48 | 0.17 | 0.5-0.67 | 0.44 | 0.48 | 0.17 | 0.0042-0.73 | 0.41 | 0.42 | 0.092 | 0.22-0.59 |
| Sum avg. (texture) peripheric ratio | 0.36 | 0.3 | 0.22 | 1.22-0.045 | 0.32 | 0.27 | 0.27 | 0.96-0.024 | 0.37 | 0.35 | 0.2 | 0.94-0.093 |
| Sum entropy (texture) peripheric ratio | 0.18 | 0.2 | 0.067 | 0.08-0.3 | 0.18 | 0.19 | 0.073 | 0.0068-0.29 | 0.17 | 0.17 | 0.049 | 0.05-0.31 |
| Sum var. (texture) peripheric ratio | 0.64 | 0.73 | 0.36 | 1.9-0.9 | 0.63 | 0.75 | 0.31 | 0.3-0.92 | 0.66 | 0.68 | 0.12 | 0.29-0.85 |
| Total energy (intensity) peripheric ratio | 0.13 | 0.015 | 0.53 | 2.93-0.54 | 0.3 | 0.027 | 0.73 | 2.23-0.43 | 0.04 | 0.073 | 0.39 | 1.33-0.4 |
| Var (intensity) peripheric ratio | 0.62 | 0.7 | 0.32 | 1.55-0.89 | 0.62 | 0.71 | 0.28 | 0.19-0.91 | 0.62 | 0.64 | 0.12 | 0.29-0.81 |

